# Reconstructing synthetic hearts from ECG using flow matching

**DOI:** 10.64898/2026.09.01.26360987

**Authors:** Jin Zheng, Soodeh Kalaie, Qiang Ma, Qingjie Meng, Khaled Rjoob, Parisa Gifani, Lei Hu, Nigar Babazade, Mattia Corianò, Wei Zhong, Majid Vafaeezadeh, Shamin Tahasildar, Nirmal Vadgama, Deva S Senevirathne, Ainkaran Santhirasekaram, Kathryn A McGurk, Lara Curran, Ying He, Long Chen, Yuanhan Mo, Ling Huang, Mengyun Qiao, Yuan Huang, Wenjia Bai, Declan P O’Regan

## Abstract

Cardiac imaging enables quantitative assessment of cardiac structure and function but remains constrained by cost, infrastructure and specialist expertise. In contrast, electrocardiogram (ECG) is widely accessible yet underexploited, despite encoding latent information about cardiac physiology. Here we introduce *visionECG*, a conditional flow matching framework that learns a probabilistic mapping between two biological distributions—the space of cardiac electrical signals and the space of cardiac geometries. Using 71,132 paired ECG and cardiac mesh sequence datasets from the UK Biobank, with external assessment in 5,000 patients with ECG-echocardiogram pairs, the model reconstructs quantitatively accurate spatiotemporal representations of the left ventricle using ECG inputs and basic demographic information alone. These reconstructions enable discrimination of structural abnormalities and disease labels, provide visualisations of functional abnormalities, and support flexible quantification of both global and regional parameters. By reframing the ECG as a generative source of patient-specific left ventricular geometry and motion, this work establishes a scalable framework for translating low-dimensional signals into high-dimensional, physiologically grounded structured representations.

## Main

The ECG is the most widely performed cardiac investigation in clinical practice, recorded in minutes, immediately interpretable, and deployable at global scale. Yet despite its ubiquity, conventional ECG interpretation is mainly limited to conduction abnormalities, leaving unexploited any latent information about cardiac structure and function encoded in waveform morphology. Cardiac diseases have a long presymptomatic course during which structural and functional abnormalities develop^1–3^, and early detection during this window could improve prognosis. Advanced cardiac imaging can detect these abnormalities but remains constrained by cost, infrastructure, and specialist expertise, limiting its use at scale^4^. Even in well-resourced health systems, imaging capacity does not scale to community screening or near-patient monitoring, and in lower-resource settings it is largely inaccessible^5,6^.

Prior work has shown that supervised deep learning applied to paired ECG and imaging data can identify ventricular dysfunction and structural heart disease with clinically useful accuracy^7–10^, translate ECG to greyscale cardiac images for disease detection^11^, and extract latent prognostic information from waveform morphology alone^12^. However, these approaches produce scalar predictions, binary classifications or unstructured data: they indicate the likely presence of pathology but do not recover an underlying representation of cardiac geometry and dynamics, which is inherently quantitative. Moving from detecting pathology to reconstructing the geometry and motion of the heart is a qualitatively different challenge—one that requires recovering high-dimensional spatial and temporal structure from a signal that captures only the integrated electrical activity of the myocardium.

Here we introduce *visionECG*, a generative model based on a conditional latent flow matching framework. *visionECG* encodes ECG waveforms and demographic variables to generate patient-specific spatiotemporal representations of left-ventricular geometry and motion throughout the cardiac cycle. Rather than producing a single prediction or disease label, the model reconstructs a structured representation from which diverse measures of cardiac morphology and performance can be derived. We assess whether these representations faithfully recover ventricular structure and mechanical function, yield quantitative phenotypes, identify structural and pathological abnormalities, and stratify cardiovascular risk. By reframing the ECG as a source of patient-specific information about cardiac morphology and dynamics, *visionECG* offers a general framework for translating routinely acquired electrical signals into structured representations of cardiac function, with the potential to extend the clinical information obtainable from the ECG beyond conventional electrocardiographic assessment.

## Results

### Study overview

We assembled an ECG–cardiovascular magnetic resonance (CMR) cohort from the UK Biobank imaging sub-study^13,14^, comprising 71,132 individuals with complete paired data (Supplementary Fig. 1; Supplementary Table 1). The cohort was partitioned at the participant level into training and test sets used consistently across all downstream tasks.

**Figure 1.**
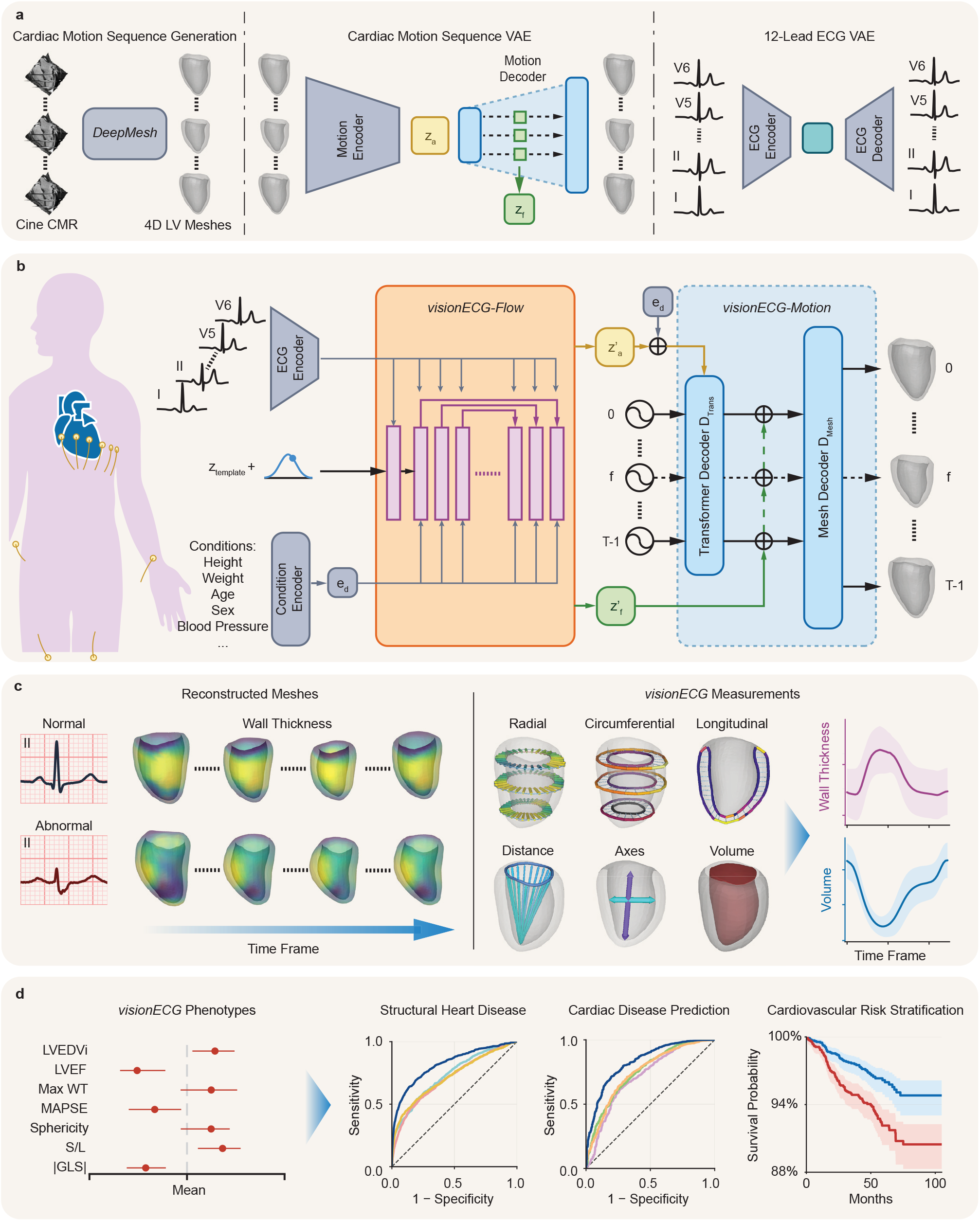
An overview of *visionECG* framework. **a**, *DeepMesh* generates four-dimensional (4D) left ventricular (LV) meshes from multiview cine CMR (short-axis and 2-chamber, 4-chamber long-axis images). VAE model pretraining: the cardiac motion sequence VAE model encodes the 4D cardiac meshes into a sequence-level latent *z*_*a*_ using a motion encoder. A motion decoder outputs frame-resolved latents *z*_*f*_ and reconstructs the cardiac mesh sequence. The ECG VAE model likewise encodes 12-lead ECG signals with an ECG encoder and reconstructs ECG waveforms with an ECG decoder. **b**, *visionECG* framework: The pretrained ECG encoder encodes ECG signals, and a condition encoder encodes demographic information into embedding *e*_*d*_ . *visionECG-Flow* takes the two encoded representations along with an anatomical prior *z*_*template*_ perturbed by Gaussian noise as inputs and generates sequence-level latent 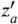 and frame-resolved latent 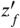. *visionECG-Motion* then combines these representations to generate the complete 4D cardiac mesh sequence. **c**, *visionECG* phenotype measurements: the global volumetric, geometric and motion-derived phenotypes are measured from a single *visionECG*-generated cardiac mesh sequence. **d**, *visionECG* clinical application: *visionECG*-derived cardiac phenotypes are compared across groups and used for cardiac disease detection and risk stratification. CMR, cardiovascular magnetic resonance; VAE, variational autoencoder; ECG, electrocardiogram; LVEDVi, left ventricular end-diastolic volume index; LVEF, left ventricular ejection fraction; WT, wall thickness; MAPSE, mitral annular plane systolic excursion; S/L, septal-to-lateral ratio; GLS, global longitudinal strain.

The ground-truth cardiac representations were generated by *DeepMesh*^15^, a multi-view CMR pipeline that integrates short-axis and long-axis cine images to reconstruct time-resolved three-dimensional (3D) left-ventricular surface meshes with vertex-wise correspondence across the cardiac cycle. By fusing orthogonal imaging planes, *DeepMesh* captures both radial and longitudinal cardiac motion, a key advantage over single-view approaches (mean surface distance error of 1.7 mm, compared to 3.0 mm for models that only use single-view imaging)^15^. For each participant, *visionECG* was trained to reproduce this output, a 50-frame mesh sequence (*1,412* vertices per frame) spanning the full cardiac cycle, from a resting 12-lead ECG and 8 demographic variables alone.

*visionECG* framework comprises two components: *visionECG-Flow* and *visionECG-Motion*, built on two pretrained variational autoencoders (VAEs) that compress the 12-lead ECG waveform and the 4D cardiac mesh sequence into compact latent representations (Fig. 1a and b). The conditional *visionECG-Flow* takes the ECG encoding, demographic variables and a stochastic anatomical prior, the population-average motion latent *z*_template_ perturbed by Gaussian noise, and generates two complementary latents: a sequence-level motion latent (*z*_*a*_) that captures cardiac motion over the complete cardiac cycle, and frame-resolved latents (*z*_*f*_) that encode the cardiac morphology at each frame. *visionECG-Motion* then combines these representations to generate the complete left-ventricular mesh sequence.

Performance was evaluated hierarchically: geometric fidelity against *DeepMesh*-derived CMR-based reference meshes; agreement with CMR-derived clinical measurements; classification of threshold-defined structural abnormalities and International Classification of Diseases (ICD)-coded cardiovascular diseases; and association with incident cardiovascular risk. To isolate the contribution of case-specific waveform information, an ECG ablation experiment replaced each participant’s ECG with a population-average waveform. Separately, to assess transferability beyond the UK Biobank, derived measurements were evaluated against echocardiographic measurements in an independent external clinical cohort.

### ECG-conditioned cardiac mesh generation

We evaluated mesh generation accuracy in 2,000 test participants using four metrics: absolute relative error in endocardial volume and myocardial wall thickness, 90^th^percentile Hausdorff distance (HD90), and average symmetric surface distance (ASSD)^16^. Since no prior ECG-to-4D-mesh generator exists, we compared *visionECG* against three adapted baselines: direct latent generation from ECG^17^; a conditional VAE^18^; and a contrastive cross-modal approach^19^, all evaluated on the same held-out participants and reference meshes.

*visionECG* outperformed all baselines across every metric at both sequence level and the end-diastolic (ED) and end-systolic (ES) frames (all paired *P* < 0.05; Fig. 2a; Supplementary Tables 2 and 3; Supplementary Fig. 2). Median absolute relative differences remained below 10% across the full sequence for both endocardial volume (8.34%) and wall thickness (9.54%). Volume error was lower at ED (median 6.91% [interquartile range (IQR) 3.30–11.65%]) than ES (8.92% [4.44–15.66%]), while wall-thickness error was lower at ES (8.27% [7.05–9.90%]) than ED (9.85% [8.23–11.69%]). Sequence-averaged surface errors were also lowest for *visionECG*: median HD90 of 3.21 mm (endocardial) and 3.37 mm (epicardial), and median ASSD of 2.06 mm and 2.19 mm, respectively.

**Figure 2.**
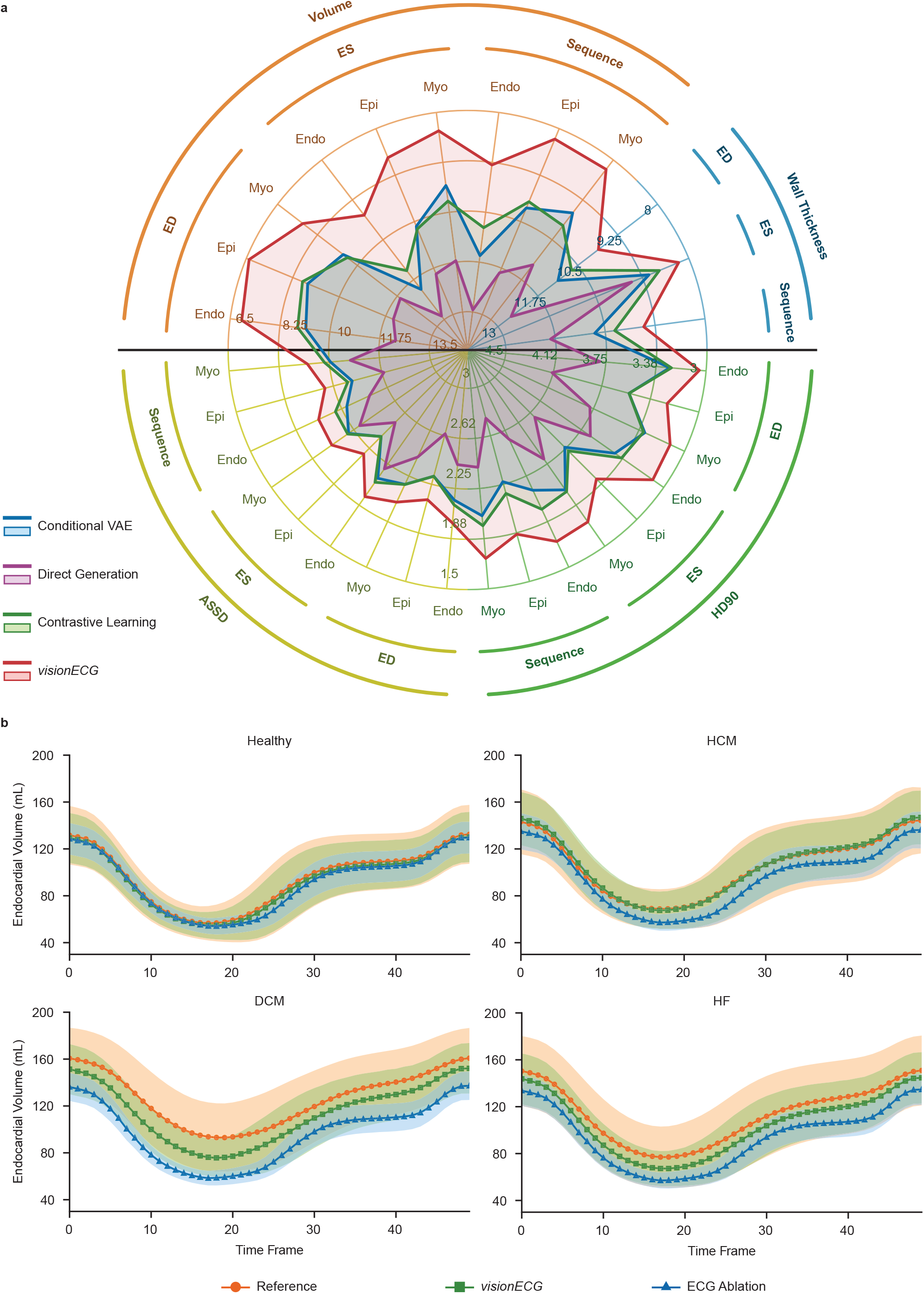
Cardiac mesh performance. a, Radar plot of cardiac mesh generation accuracy: cardiac mesh generation accuracy of *visionECG* is compared against other benchmarks using four metrics: absolute relative errors in volume and wall thickness, HD90 and ASSD, relative to reference meshes. Metrics are evaluated at ED and ES frames and across the full sequence, for endocardial, epicardial and myocardial meshes where applicable. **b**, Population-average ECG ablation: participant-specific ECG are replaced with population-average waveforms, then endocardial volume trajectories from the ECG ablation model are compared with reference and *visionECG* across healthy participants and disease groups. HD90, 90^th^percentile Hausdorff distance; ASSD, average symmetric surface distance; ED, end diastole; ES, end systole; Endo, endocardial; Epi, epicardial; Myo, myocardial; VAE, variational autoencoder; ECG, electrocardiogram; HCM, hypertrophic cardiomyopathy; DCM, dilated cardiomyopathy; HF, heart failure.

To confirm that *visionECG-Flow* exploits waveform information beyond demographic priors, we replaced each participant’s ECG with a population-average waveform across healthy, hypertrophic cardiomyopathy (HCM), dilated cardiomyopathy (DCM) and heart failure (HF) groups (total *n* = 5,253; Fig. 2b; Supplementary Tables 1 and 4). ECG removal increased median absolute relative volume error from 7.64% [4.54–12.32%] to 10.33% [6.41–16.55%] in healthy participants (*P* < 10^−16^), with substantially larger degradation in disease groups: 10.25% to 14.86% in HCM, 12.59% to 20.29% in DCM, and 10.65% to 16.97% in HF (all *P* ≤ 0.0099). Without the case-specific ECG, generated volume trajectories collapsed toward the population mean, losing the characteristic asymmetry of the true cardiac cycle—confirming that ECG waveform encoding is especially critical for reproducing abnormal cardiac motion.

### Synthetic meshes accurately recover cardiac phenotypes

Because all generated meshes share anatomical correspondence through *visionECG-Motion*, a diverse panel of imaging-derived measurements can be extracted from a single synthetic sequence without retraining. From each *visionECG*-generated sequence we derived global volumetric phenotypes (end-diastolic volume (EDV), end-systolic volume (ESV), left ventricular mass (LVM), left ventricular ejection fraction (LVEF)), geometric phenotypes (maximum wall thickness (WT), segmental wall thickness, sphericity index, septal-to-lateral ratio) and motion-derived phenotypes (mitral annular plane systolic excursion (MAPSE), global circumferential strain (GCS), global longitudinal strain (GLS)) (Fig. 3 and Supplementary movie 1).

**Figure 3.**
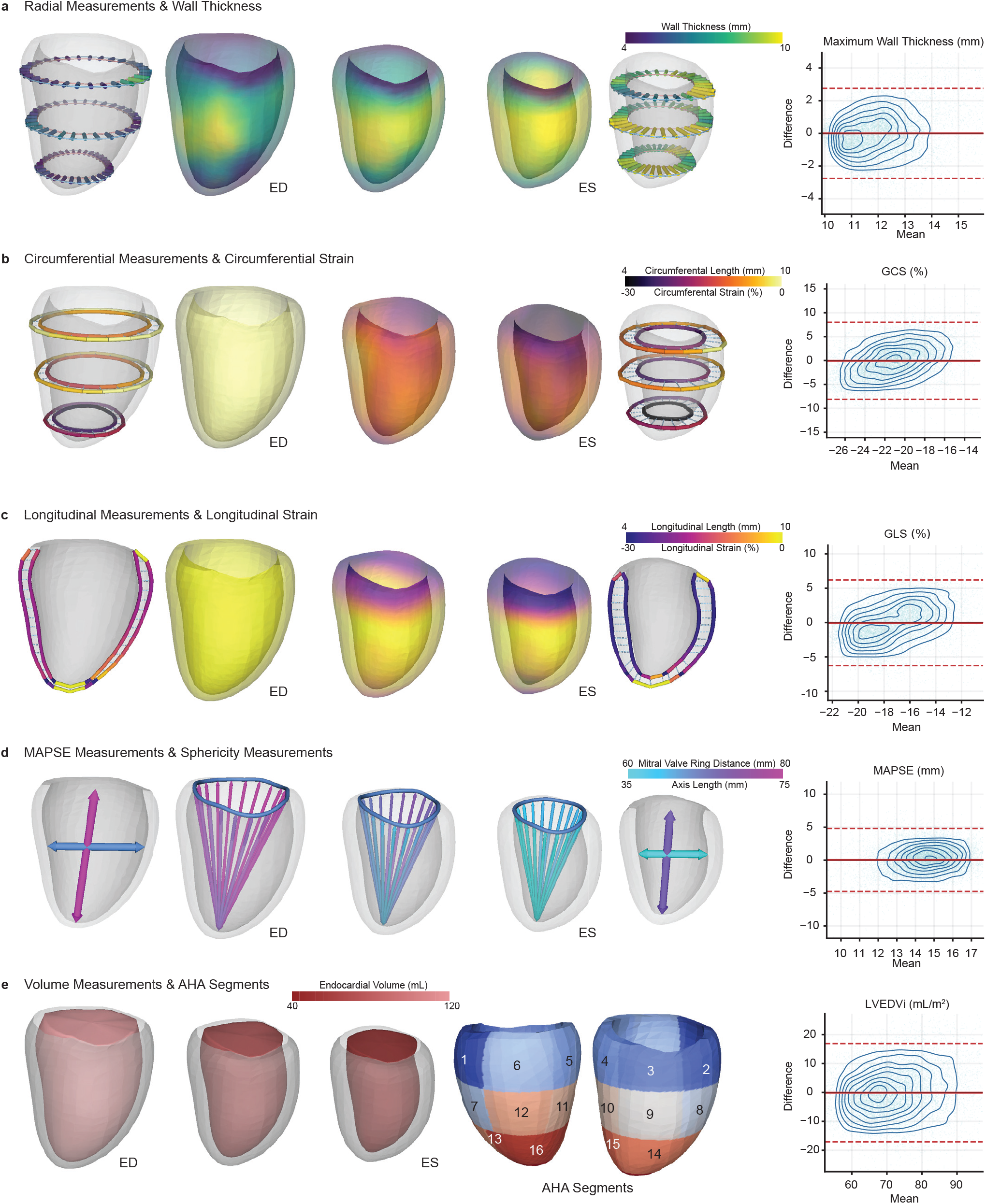
Visualisation and validation of *visionECG*-derived cardiac phenotype measurements. **a-e**, Measurements of cardiac volumetric, geometric and motion-derived phenotypes are visualised on the *visionECG*-generated cardiac mesh sequence, from end diastole to end systole. **a**, radial measurements and wall thickness; **b**, circumferential measurements and circumferential strain; **c**, longitudinal measurements and longitudinal strain; **d**, MAPSE and sphericity measurements; **e**, volume measurements. The Bland-Altman plots (right) assess agreement between *visionECG*- derived measurements and reference CMR-derived measurements. For each plot, the solid line is the mean bias; the dashed lines are the 95% limits of agreement. Detailed values are provided in Supplementary Table 5. ED, end diastole; ES, end systole; GCS, global circumferential strain; GLS, global longitudinal strain; MAPSE, mitral annular plane systolic excursion; LVEDVi, left ventricular end-diastolic volume index; AHA, American Heart Association.

Bland–Altman analysis of the full test set (*n* = 14,226) showed good agreement between *visionECG*-derived and CMR-derived measurements across all phenotype classes, with no significant bias (Fig. 3, Supplementary Fig. 3, Supplementary Table 5). The generated sequences reproduced smooth cardiac-cycle trajectories for endocardial volume, epicardial volume and wall thickness, confirming that the temporal structure encoded in both sequence-level latent (*z*_*a*_) and frame-resolved latents (*z*_*f*_) is decoded across the full cycle. Supplementary Fig. 4 illustrates an example of the association between the ECG morphologies and *visionECG* phenotypes, with taller precordial R and T waves positively associated with LVM.

**Figure 4.**
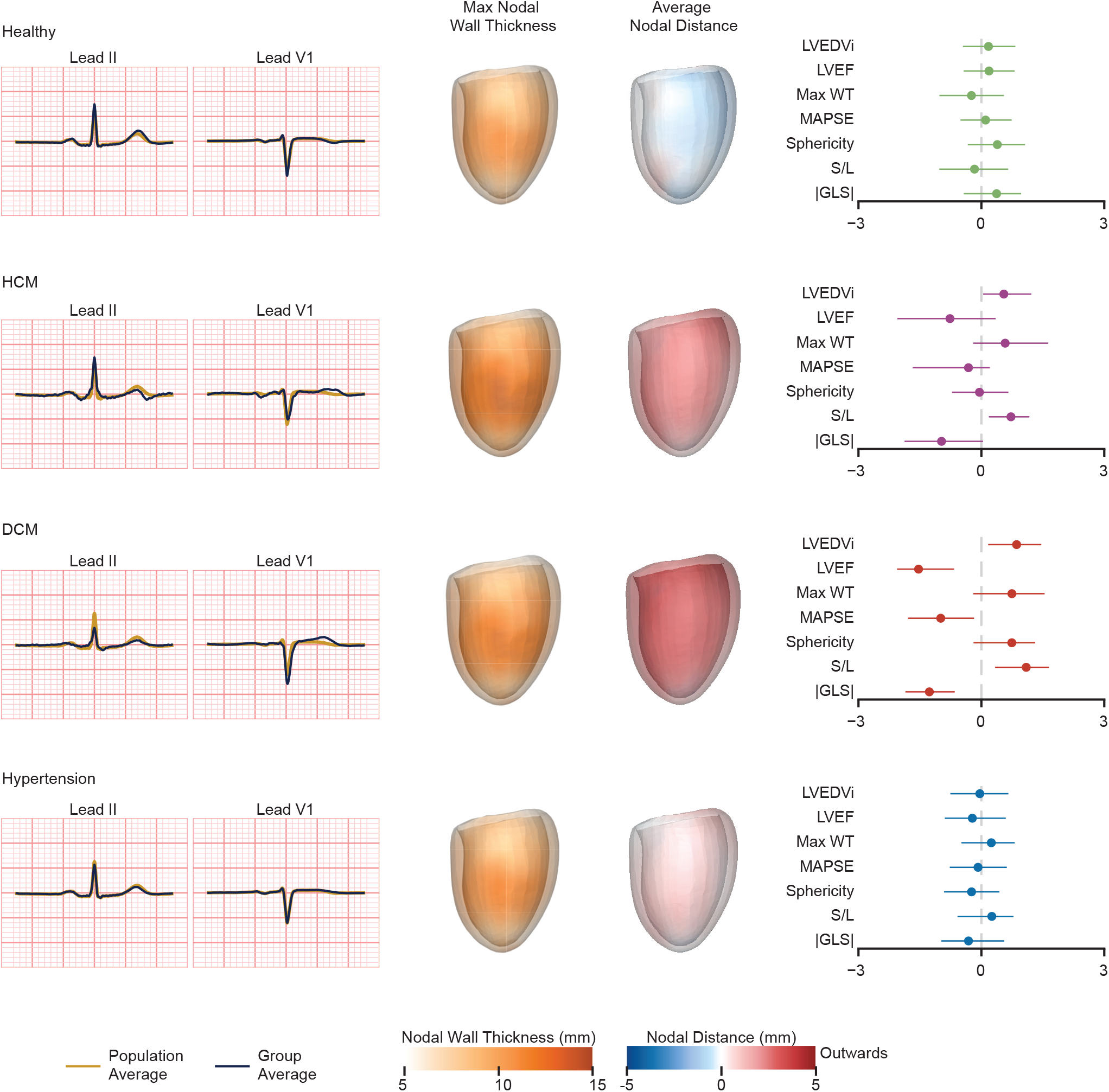
Group-specific cardiac phenotypes. Left, population-average and group-average ECG waveforms. Middle, maximum nodal wall thickness and average nodal distance mapped onto cardiac meshes at the end-diastolic frame. Average nodal distance represents the mean distance of each mesh node relative to the population-average mesh. Red and blue colours indicate outward and inward distance, respectively. Right, forest plots summarise cardiac phenotype profiles across groups, normalised using the Yeo-Johnson method. Points and error bars indicate normalised medians and interquartile ranges. HCM, hypertrophic cardiomyopathy; DCM, dilated cardiomyopathy; LVEDVi, left ventricular end-diastolic volume index; LVEF, left ventricular ejection fraction; WT, wall thickness; MAPSE, mitral annular plane systolic excursion; S/L, septal-to-lateral ratio; GLS, global longitudinal strain.

The generated meshes preserved disease-associated phenotype profiles across healthy, HCM, DCM and hypertensive participants (Fig. 4; Supplementary Fig. 5). Relative to healthy participants, HCM showed greater maximum WT (12.70 [11.51–15.18] mm vs. 11.45 [10.56–12.64] mm; adjusted *P* = 2.5 × 10^−10^); DCM showed ventricular dilatation and markedly lower LVEF (44.20 [29.92–53.71]% vs. 60.29 [55.66–64.35]%; *P* < 1× 10^−16^); and hypertension was associated with increased maximum WT (12.15 [11.15–13.15] mm; *P* < 1 × 10^−16^). Lower wall thickness values are obtained by 3D analysis, which assesses true orthogonal measurements without the in-plane obliquity of conventional imaging^20^. Full pairwise comparisons are in Supplementary Table 6.

**Figure 5.**
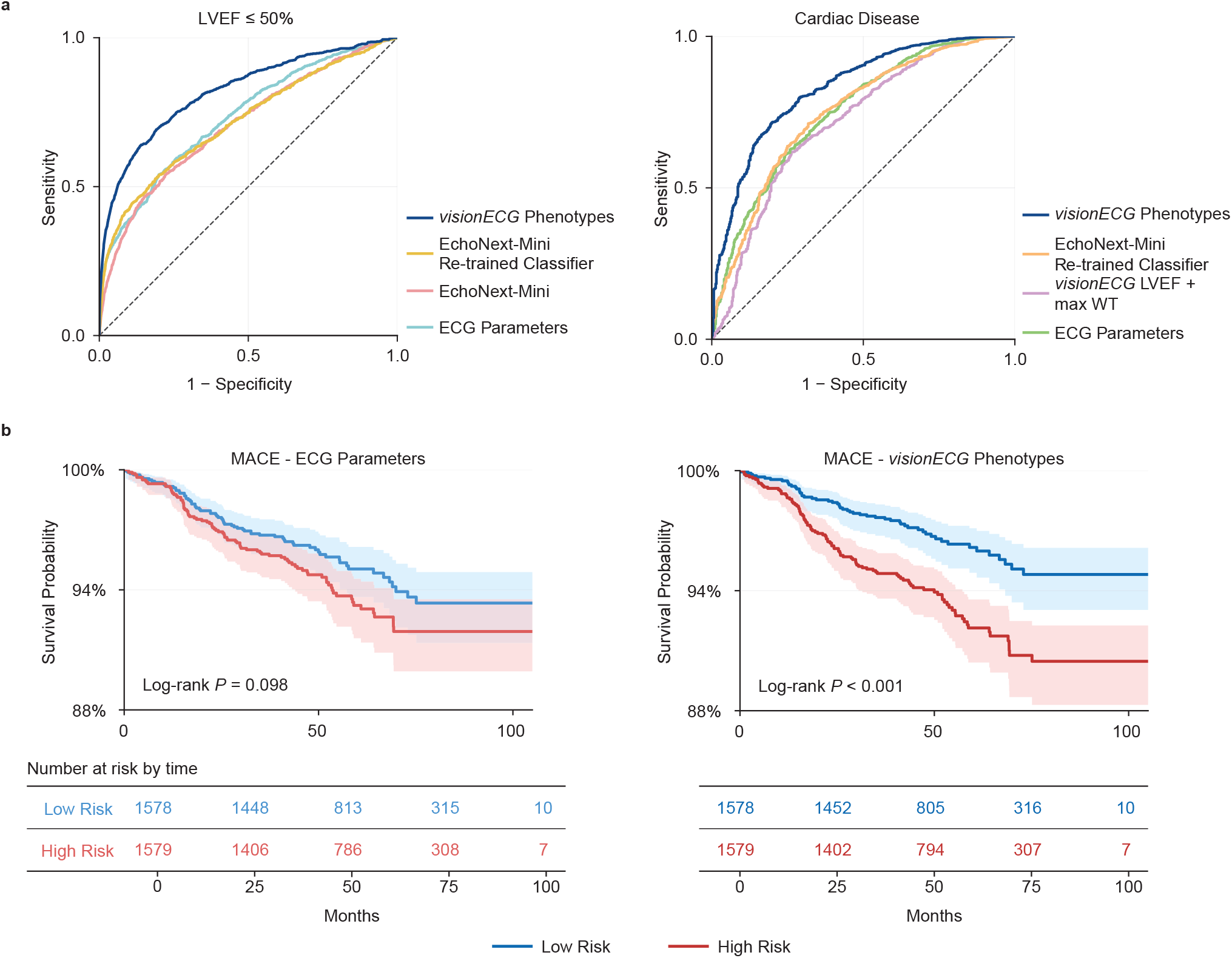
Disease discrimination and outcome prediction. **a**, Receiver operating characteristic curves for LVEF≤ 50% and overall cardiac disease comparing *visionECG* to other benchmarks. Area under the receiver operating characteristic curve with 95% confidence intervals (CI) refers to Supplementary Tables 8 and 10. **b**, Kaplan-Meier plots for *visionECG* vs. conventional ECG parameters. For both models, participants are divided into low- and high-risk groups by median risk score. For each plot, the Log-rank test is performed to compare survival between risk groups. Hazard ratios with 95% CIs refer to Supplementary Table 12. LVEF, left ventricular ejection fraction; MACE, major adverse cardiovascular events.

Importantly, although these phenotypes were not included as explicit training targets, they could still be computed post hoc from the same generated mesh sequence. Computing LVM, MAPSE, GCS, GLS, sphericity index, septal-to-lateral ratio, and 17-segment wall thickness revealed complementary disease signatures: hypertension combined increased LVM with reduced absolute GLS; DCM showed decreased absolute GCS and greater sphericity; and HCM exhibited a higher septal-to-lateral ratio and more impaired GLS than hypertension (all adjusted *P* < 0.01; Fig. 4; Supplementary Fig. 5; Supplementary Table 7). Examples of disease cases and the healthy average are provided in Supplementary Fig. 6 and Supplementary movie 2. A single generated mesh sequence thus serves as a reusable substrate from which arbitrary geometric and motion phenotypes can be derived on demand.

### Interpretable detection of structural heart disease across different settings

We next tested whether phenotype measurements from the same *visionECG*-generated mesh sequences could identify clinically relevant structural abnormalities. We evaluated two complementary tasks. First, we applied imaging-based thresholds to left ventricular end-diastolic volume index (LVEDVi), LVEF and maximum WT to define left-ventricular dilatation, systolic dysfunction and increased wall thickness. Second, we tested whether the full set of *visionECG*-derived phenotypes could classify ICD code-based major cardiac diseases, including HF, myocardial infarction (MI), cardiomyopathy (CM), HCM and DCM, as well as overall major cardiac disease, defined as the presence of any of these conditions.

For threshold-defined abnormalities, *visionECG* achieved area under the receiver operating characteristic curve (AUC)≥ 0.80 on all six tasks (AUCs=0.80 for maximum WT≥ 13 mm and for LVEF≤ 45%), while all reference models—least absolute shrinkage and selection operator (LASSO) models of conventional ECG parameters and EchoNext-Mini-based comparators^9,21^—fell below AUC 0.76 on every task (two-sided DeLong, *P* < 0.05; Fig. 5a; Supplementary Fig. 7; Supplementary Table 8). Sensitivity was ≥0.58 and specificity was ≥0.72 across all tasks.

To assess transferability to echocardiographic measurements and generalisability to an independent clinical population, we evaluated *visionECG* in an age-matched external cohort (*n* = 5,000) comprising paired 12-lead ECG and echocardiogram from a tertiary hospital^21,22^. The LVEF derived from the *visionECG*-generated meshes achieved AUC values of 0.79, 0.80 and 0.81 for detecting echocardiographic LVEF ≤ 50%, ≤ 45% and ≤ 35%, respectively (Supplementary Table 9).

For ICD-coded diseases (*n* = 18,273), the *visionECG* extreme gradient boosting (XGBoost) model achieved the highest discrimination for overall cardiac disease (AUC 0.83 [95% confidence interval (CI) 0.81, 0.85]), HF (0.82 [0.78, 0.86]) and MI (0.81 [0.79, 0.83]), outperforming both comparators (all *P* < 0.05; Fig. 5a; Supplementary Fig. 8; Supplementary Table 10). HCM classification improved significantly over both comparators (*P* < 0.05), while DCM performance was not significantly different from the retrained EchoNext-Mini comparator. These results demonstrate that the structured phenotype panel derived from *visionECG* mesh sequences enables interpretable, multi-disease detection that consistently exceeds waveform-only approaches.

### ECG-derived phenotypes independently stratify incident cardiovascular risk

In the 17,566 participants with follow-up records, XGBoost-derived 5-year prognostic indices from either conventional ECG parameters or *visionECG* phenotypes were entered into Cox proportional hazards (PH) models for incident HF, MI and major adverse cardiovascular events (MACE) (Supplementary Table 11).

*visionECG* phenotypes discriminated risk more effectively than ECG parameters across all three outcomes, with the largest advantage for HF (Harrell’s concordance index (C-index) 0.76 [0.70, 0.82] vs. 0.63 [0.54, 0.71]; Supplementary Table 12 and corresponding Kaplan–Meier plots (Fig. 5b; Supplementary Fig. 9)). The *visionECG* phenotype index was a significant independent predictor of every outcome (for example, hazard ratio 1.79 [1.60, 2.00] per standard deviation (SD) for HF; all *P* < 0.05), whereas the ECG parameter index was not significant for MI. In joint models, adding *visionECG* phenotypes to ECG parameters significantly improved discrimination for all three outcomes (all *P* < 0.05), whereas the reverse addition did not (all *P* > 0.05; Supplementary Table 13). Together, these findings establish that *visionECG* phenotypes carry independent prognostic information that is not captured by conventional waveform measurements alone.

## Discussion

Conventional ECG interpretation extracts a small fraction of the physiological information encoded in cardiac electrical activity^8^. Even contemporary deep learning applied to ECG signals produces only scalar predictions or binary classifications, indicating likely pathology without recovering the underlying cardiac representation^7,9,12^. Here we show how *visionECG* uses flow matching to learn a principled probabilistic bridge between two biological distributions—the space of cardiac electrical signals and the space of cardiac geometries. *visionECG* transports a stochastic anatomical prior through a motion-latent space conditioned on ECG and demographic inputs, generating a sequence-level latent that captures subject-level cardiac identity and frame-resolved latents that encode beat-by-beat dynamics. Unlike VAE or generative adversarial network (GAN)-based approaches^18,23^, flow matching provides stable training and tractable likelihoods across high-dimensional, temporally ordered outputs. The factored latent structure captures both overall cardiac shape and beat-by-beat motion within a single compact representation, decoded into a full mesh sequence in a single forward pass. As the output is an anatomically consistent, correspondence-preserving mesh sequence, arbitrary phenotypes–volumetric, geometric, strain-based and segmental—can be derived post hoc without retraining, including measurements not defined at the time of model training.

Disease-associated patterns of hypertrophy, dilatation and contractile impairment were preserved across groups, and the full phenotype panel consistently outperformed waveform-only models across disease detection and risk stratification tasks. That these structured cardiac representations outperform direct ECG-to-label mappings suggests that recovering the physiological substrate, rather than predicting summary statistics, is the more powerful and generalisable strategy—one that becomes more valuable as new phenotypes are defined, without requiring the model to be retrained. These properties open a potential role for *visionECG* in settings where imaging is unavailable or impractical. Pre-referral triage in primary care, opportunistic screening in community settings and longitudinal monitoring from serial ECGs are all plausible future applications, particularly in lower-resource settings where the gap between ECG availability and imaging access is largest^4^. Differentiating between hypertension and HCM, which both have increased mass but with different morphologies, is an example of where a phenotypic reconstruction is an advantage over tabular data alone.

There are several limitations to consider. The model was trained and validated within the UK Biobank, a volunteer cohort with predominantly European ancestry. Although we performed external assessment in a hospital cohort, further evaluation in more diverse ancestry groups and clinical settings, and fine-tuning for different use cases, will be needed to better assess model portability and support clinical utility. The generated meshes approximate, but do not replace, structural assessment, with residual geometric errors that were greater at end-systole, where the ECG provides less direct information about ventricular configuration. The model also tended to underestimate more localised spatial features relative to global changes, which are likely to be better represented in the ECG signal. In addition, echocardiography and CMR provide information beyond ventricular morphology and function, including valve assessment, tissue characterization and full four-chamber evaluation. Nonetheless, the generated meshes retain practical value as structured left-ventricular representations from which multiple specific measures, including strain, regional wall thickness and shape indices, can be computed from the same model.

The broader implication of this work is a reframing of what low-dimensional biosignals can provide. Rather than optimising for any single prediction target, this approach treats the structured dynamic representation as the primary output—one that is interpretable, reusable, and extensible across measurements and disease contexts. Phenotyping, detection and prognosis then follow from the same generated representation without additional supervision. Extending this paradigm to other routinely acquired signals, including ambulatory and wearable ECG recordings, could enable longitudinal tracking of cardiac remodelling at population scale, establishing a new class of cardiac assessment that is simultaneously quantitative, generalisable and accessible.

## Methods

### UK Biobank participants

UK Biobank is a large cohort study of half a million individuals in the UK aged 40-69 at recruitment between 2006 and 2010^24^. All participants provided written informed consent for the study, which was also approved by the National Research Ethics Service (11/NW/0382). Our study was conducted under terms of access approval number 40616. A range of phenotypic and genotypic attributes, risk factors and physical measures were used for the analyses. These were collected by touchscreen questionnaires, interviews, biophysical measurements, hospital episode statistics, and primary care data. CMR images were also acquired for a subset of participants. Details of how each phenotype was acquired are available on the UK Biobank Showcase (http://biobank.ctsu.ox.ac.uk/crystal/).

### 4D motion extraction

CMR imaging was performed on participants to capture two-dimensional (2D) retrospectively gated cine imaging on a 1.5T magnet (Siemens Healthineers, Erlangen, Germany)^25^. Left ventricular short-axis plane cine images from base to apex were acquired, as well as long-axis cine images in the two- and four-chamber views. Cines comprised 50 cardiac frames with a typical temporal resolution of 31 ms. Ascending and descending thoracic aorta images were also acquired with transverse cine imaging.

The left ventricle is represented as a 4D surface mesh that includes both the endocardial and epicardial surfaces. We adopt a deep learning framework (*DeepMesh*^15^) to estimate 4D cardiac motion on this mesh from multi-view 2D cine CMR images, including short-axis stacks and long-axis views (2-chamber and 4-chamber, Fig. 1a). Unlike traditional image-space methods that estimate dense pixel-or voxel-level displacements, *DeepMesh* operates directly in mesh space and learns per-vertex displacements, preserving one-to-one vertex correspondence across time and subjects. This mesh-based formulation avoids interpolation artefacts when projecting image-based motion fields onto anatomy, simplifies population-level analysis, and enables direct computation of clinically relevant volumetric and regional functional biomarkers on anatomically consistent meshes.

The *DeepMesh* framework consists of two stages: (i) Reconstruction of a subject-specific end-diastolic mesh from a population template; and (ii) Estimation of per-vertex motion across the cardiac cycle by deforming the end-diastolic mesh over time. Both stages are supervised using a differentiable rasteriser that projects the evolving 3D mesh into soft 2D contours on short-axis and long-axis planes, enabling the use of standard 2D annotations for training. The inclusion of long-axis views in this supervision is critical to improving motion estimation along the heart’s long axis. In the first stage, a subject-specific end-diastolic mesh is reconstructed by deforming a fixed-topology population template mesh. A convolutional neural network processes the multi-view end-diastolic images to predict a 3D voxel-wise displacement field. This field is sampled at the template’s vertex locations to produce the deformed mesh for that subject. As a result, all end-diastolic meshes are topologically consistent (i.e., share the same vertex and face structure) and directly comparable across individuals.

In the second stage, *DeepMesh* estimates the left ventricle motion over time by predicting per-vertex displacements from the end-diastolic frame to any time point t in the cardiac cycle. A motion network processes the multi-view images at the end-diastolic frame and frame t, and predicts a 3D voxel-wise motion field ϕ_0→*t*_ that captures how the myocardium deforms over time. Sampling this field at the end-diastolic mesh vertex locations yields the per-vertex displacement Δ*V*_0→*t*_, which is used to deform the end-diastolic mesh to the mesh at time 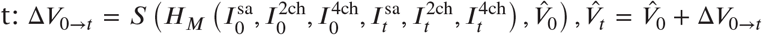. Here, *H*_*M*_ (⋅) denotes the motion network, which outputs the intermediate motion field ϕ_0→*t*_, and *S*(⋅) samples it at the mesh vertices. This results in a time-resolved sequence of anatomically consistent meshes that capture dynamic left ventricle motion across the entire cardiac cycle.

To supervise training using standard 2D annotations, *DeepMesh* employs a differentiable rasteriser that projects 3D meshes into 2D probability maps representing myocardial contours. The rasteriser slices the predicted mesh along the short-axis and long-axis planes and computes each vertex’s likelihood of lying on the image plane based on its distance to the plane. The resulting 2D soft contours are then compared to ground-truth 2D annotations using a weighted Hausdorff distance, a robust loss function that handles sparse or noisy annotations. This rasterisation strategy provides strong supervision from all three anatomical views. In particular, the short-axis view contributes dense in-plane anatomical coverage across slices, while the long-axis views provide high-resolution through-plane constraints that are critical for accurate reconstruction and motion estimation along the long axis.

In addition to the contour-based loss, model training incorporates auxiliary constraints to ensure anatomical plausibility and motion coherence. Laplacian smoothness promotes surface regularity, Huber loss encourages coherent motion fields, and an image similarity loss provides self-supervision by enforcing consistency between the warped and reference short-axis stack. All losses are fully differentiable and jointly optimised in an end-to-end manner, enabling robust learning from multi-view cine CMR using only standard 2D annotations.

### Mesh-based phenotype measurements

All phenotypes were measured directly from the generated cardiac motion sequence, whose endocardial and epicardial vertices are in one-to-one correspondence across frames and subjects. The phenotypes can be clustered into global volumetric phenotypes, including EDV, ESV, LVM, and LVEF; geometric phenotypes, including global WT, segmental WT, sphericity index and septal-to-lateral wall thickness ratio; and motion-derived phenotypes, including MAPSE, GCS and GLS (Fig. 3 and Supplementary movie 1). At each frame, the volumes enclosed by the endocardial and epicardial vertex sets were calculated by closing the open basal ring of each surface and integrating the enclosed volume^26^. Myocardial volume was computed as the epicardial minus the endocardial volume, and LVM was obtained by multiplying by a myocardial density of 1.05 g mL^−1^. LVEF was calculated as (LVEDV − LVESV)/LVEDV × 100%. Volumes and mass were indexed to body surface area (BSA) using the Du Bois formula: BSA = 0.007184 × Height^0.725^ × Weight^0.425^, with height in cm and weight in kg. The wall thickness was measured as the distance between the epicardium and endocardium. The maximum WT was the maximum value across all vertices at the ED frame. The sphericity index is the ratio of the short-axis and long-axis diameters of the volume enclosed by the endocardium computed by principal-component analysis of the endocardial vertex cloud^27,28^. The left ventricle was divided into 17 standardised anatomical segments for segmental measurements^29^. MAPSE was measured by longitudinal displacement of the mitral ring, median reduction in all pairwise distances between mitral-annular vertices and the apical endocardial vertex, from ED to ES frame^30^. Circumferential and longitudinal strains were calculated by 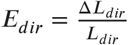 with *dir* the circumferential or longitudinal direction, *L*_*dir*_ the length of a line segment in this direction and Δ*L*_*dir*_the change of length over time^31^. GCS and GLS were reported as the global peak strains. Secondary measurements were linearly calibrated in the training set using the least-squares method.

### ECG parameters measurements

Resting 12-lead ECG was acquired from UK Biobank participants during the imaging assessment visit (data field 20205) at a sampling rate of 500 Hz, comprising the standard lead set (I, II, III, aVR, aVL, aVF, V1–V6). Recordings with missing leads or failing automated quality checks were excluded.

Raw ECG signals were preprocessed first by removing baseline wander and high-frequency noise. R-peaks were then detected in Lead II, and a 1.2-second window (0.6 s before and 0.6 s after each R-peak) was extracted across all 12 leads at 500 Hz, with the median across all detected beats yielding a single representative waveform per lead. Finally, the per-lead mean was subtracted to remove residual DC offset.

Scalar ECG phenotypes were derived from automated UK Biobank measurements, including heart rate, PR interval, QRS duration, QT interval, rate-corrected QT (QTc), P-wave axis, QRS axis, and T-wave axis.

### Problem formulation and latent rectified flow matching

#### Problem formulation

*visionECG* generates a 4D cardiac motion sequence 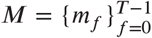 from a 12-lead ECG and basic demographic information, where each *m*_*f*_ is a triangulated left-ventricular surface mesh at cardiac frame *f* . All meshes share a common topology across frames and subjects. The conditioning inputs (Fig. 1b) are the raw 12-lead ECG *K*_ecg_ ∈ ℝ^12×*L*^of length *L*, and a demographic vector *K*_demo_ ∈ ℝ^8^comprising age, sex, height, weight, body mass index, body surface area, and systolic and diastolic blood pressure. The learning objective is therefore the conditional distribution *p*(*M* ∣ *K*_ecg_, *K*_demo_).

To make this high-dimensional generative problem tractable, *visionECG* performs conditional flow matching in compact latent spaces of the ECG and the cardiac motion sequence (Fig. 1a). It comprises two components (Fig. 1b): *visionECG-Flow*, which synthesises motion latents conditioned on the encoded ECG signals and demographic variables, and *visionECG-Motion*, which fuses these latents and generates a fully resolved 4D cardiac motion sequence.

#### Latent representations

The two latent spaces are learned by a 12-lead ECG VAE and a cardiac motion sequence VAE, respectively (Fig. 1a). The ECG VAE^17,32^is a dilated convolutional variational autoencoder for 12-lead resting ECG. The cardiac motion sequence VAE^18^couples mesh-aware graph convolutions within each cardiac phase with a temporal Transformer that fuses vertex features across the cardiac cycle.

The cardiac motion sequence VAE provides two complementary representations of the same motion sequence at different temporal granularities. The sequence-level latent *z*_*a*_ ∈ ℝ^*d*^, i.e., the memory token of the encoder Transformer, compresses the whole cycle into a single vector and captures its global spatiotemporal dynamics. The frame-resolved latents 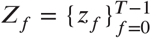, *z*_*f*_ ∈ ℝ^*d*^, taken at the output of the decoder Transformer before mesh decoding, provide an explicit frame-specific representation for reconstructing the mesh at each cardiac phase.

#### Latent rectified flow matching objective

Flow matching^33^learns a time-dependent vector field *v*_*θ*_, with neural network parameters *θ*, that defines a continuous normalising flow

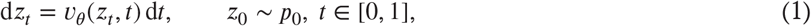

transporting a source distribution *p*_0_ = *p* to a data distribution *p*_1_ = *q* along a probability path {*p*_*t*_}_*t*∈[0,1]_ with *z*_*t*_ ∼ *p*_*t*_. Direct regression to the marginal vector field generating this path is intractable, so the flow is learned by the conditional flow matching loss

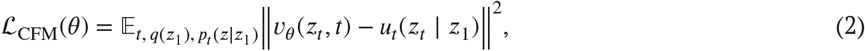

where the conditional probability path *p*_*t*_(*z* ∣ *z*_1_) satisfies ∫ *p*_*t*_(*z* ∣ *z*_1_) *q*(*z*_1_) d*z*_1_ = *p*_*t*_(*z*) with a corresponding conditional vector field *u*_*t*_(*z* ∣ *z*_1_)^33^. Following the rectified-flow construction^34,35^, we adopt the linear interpolation of *z*_0_ and *z*_1_,

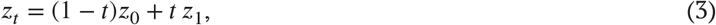

for which the conditional vector field reduces to the constant straight path *u*_*t*_(*z*_*t*_) = *z*_1_ − *z*_0_.

Given the complexity of the 4D cardiac motion sequence, we train this flow in the motion latent space, so that Eq. (1) becomes

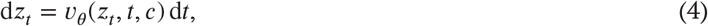

where *c* is the conditioning bundle carrying the ECG, demographic and frame-condition embeddings, and the target is *z*_1_ = *z*_*a*_ or *z*_1_ = *z*_*f*_ . Since *z*_*a*_ and *z*_*f*_ share the same dimension, a single network synthesises both granularities, selected by a frame condition *i* ∈ {0, 1, …, *T*}: *i* = 0 requests the sequence-level latent *z*_*a*_, and *i* = *f* + 1 requests the frame-resolved latent *z*_*f*_ . The source *z*_0_ is drawn from a stochastic anatomical prior defined directly in the motion latent space,

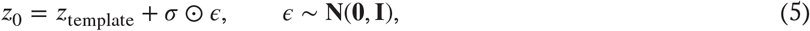

where *z*_template_ and σ are the empirical mean and per-dimension standard deviation of the training motion latents. This anchors every generative trajectory at the population-average cardiac shape and motion while preserving the stochasticity required to model subject-specific variability. Eq. (2) then specialises to the latent rectified flow matching loss

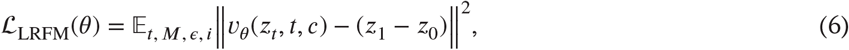

with the expectation taken over *t* ∈ [0, 1], *M* ∼ *q*(*M*), ϵ ∼ **N**(**0, 1**) (which determines *z*_0_) and *i* ∈ {0, 1, …, *T*} (which determines *z*_1_). In short, the flow network maps (*z*_0_, *K*_ecg_, *K*_demo_) to generated motion latents 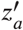 and 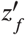 trained towards the targets *z*_*a*_ and *z*_*f*_ encoded from the observed 4D cardiac motion sequence.

At inference, we draw *z*_0_ from the stochastic prior and integrate Eq. (4) with *N* uniformly spaced explicit Euler steps, 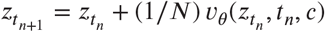.

### Model architecture and training

***visionECG* framework**. *visionECG*, composed of *visionECG-Flow* and *visionECG-Motion*, transports a stochastic anatomical prior through a motion-latent space conditioned on the encoded ECG signals and demographic variables and then generates a 4D cardiac motion sequence for interpretable assessment of cardiac structure and function.

#### visionECG-Flow

*visionECG-Flow* generates complementary sequence-level and frame-resolved motion latents. It transports a stochastic, population-informed prior through the cardiac-motion latent space to generate the sequence-level latent *z*_*a*_ and the frame-resolved latent *z*_*f*_, conditioned on the encoded ECG signals, participant demographics and frame condition. A conditional U-Net parameterises the time-dependent velocity field *v*_*θ*_(*z*_*t*_, *t, c*) of Eq. (4), where *t* is the flow time and *c* is the conditioning bundle defined below^36^.

Each conditioning input is projected by a dedicated encoder into the conditioning bundle *c* = (*e*_ecg_, *e*_d_, *e*_frame_). The ECG embedding *e*_ecg_ is obtained by passing the 12-lead ECG through the frozen ECG VAE encoder followed by an multilayer perceptron (MLP); the demographic embedding *e*_d_ and the frame embedding *e*_frame_ are produced by two further MLPs. Conditioning starts by concatenating the ECG embedding with *z*_*t*_ and linearly projecting to the U-Net hidden width, while the remaining conditions are injected at every residual block through adaptive layer normalisation (AdaLN)^37^,

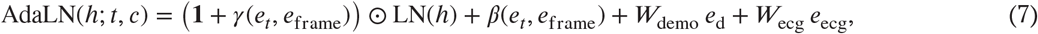

where *h* is the input feature, *e*_*t*_ a learned flow-time embedding, γ and β linear maps producing element-wise scale and shift, and *W*_demo_, *W*_ecg_ block-specific linear projections. This routing sends the trajectory-dependent signals (*t, i*) through the scale-and-shift path, while the subject-specific signals (*e*_d_, *e*_ecg_) modulate the feature space additively.

#### visionECG-Motion

*visionECG-Motion* fuses the generated sequence-level latent 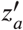 with the frame-resolved latent sequence 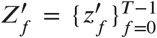, integrating global cardiac-cycle context with phase-specific geometry, and decodes them into the complete 4D cardiac motion sequence. To couple the two latent streams and promote temporal coherence across the cardiac cycle, we adapt the pretrained 4D motion mesh decoder by inserting residual modules before the Transformer decoder *D*_Trans_ and before the mesh decoder *D*_Mesh_. The generated 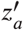, concatenated with *e*_d_ to provide subject-specific context, passes through the first module and serves as the cross-attention memory of *D*_Trans_, which frame tokens query to produce a per-frame latent sequence 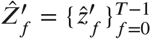 consistent with the sequence-level representation. Each *z*^′*f*^is then concatenated with the matched 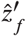, passed through the residual modules and decoded by *D*_Mesh_ to yield the 4D cardiac motion sequence *M*.

#### Training

Training of the *visionECG* framework proceeds sequentially. We first pretrain the 12-lead ECG VAE and cardiac motion sequence VAE for latent representation; then train *visionECG-Flow* to generate motion latents in the cardiac-motion latent space, conditioned on the encoded ECG signals and demographic variables; and finally adapt *visionECG-Motion* to generate complete cardiac motion sequences from the flow-generated latents.

The two VAEs are pretrained independently. The ECG VAE uses a 1024-dimensional latent representation and an input length of *L* = 600 samples, and is trained using a reconstruction mean squared error (MSE) loss and a Kullback–Leibler (KL)-divergence loss,

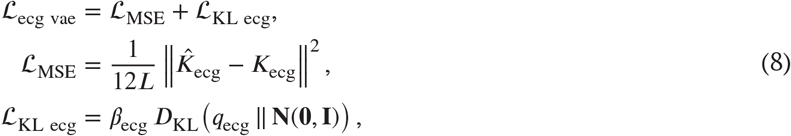

where 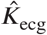 and *K*_ecg_ are the reconstructed and ground-truth ECG signals, and *D*_KL_ encourages the approximate posterior *q*_ecg_, the latent distribution predicted by the encoder for each input ECG, to stay close to the standard Gaussian prior **N**(**0, 1**), weighted by β_ecg_. The ECG VAE is trained with the Adam optimiser (learning rate 10^−4^, KL weight 0.01, 300 epochs).

The cardiac motion sequence VAE uses a 512-dimensional latent representation and is trained with Chamfer-distance reconstruction^38^, Laplacian mesh-smoothing^39^and KL-divergence losses,

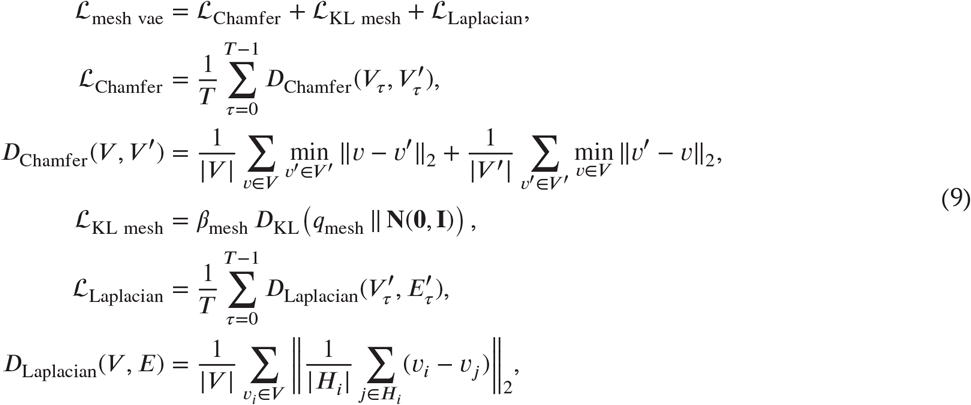

where *V* and *V* ^′^are the ground-truth and reconstructed mesh vertex coordinates, *q*_mesh_ is the approximate posterior over the motion latent with KL weight β_mesh_, and *H*_*i*_ is the set of vertices adjacent to *v*_*i*_ under the edge set *E*. The cardiac motion sequence VAE is trained with the Adam optimiser (learning rate 10^−4^, KL weight 0.01, 300 epochs)^18^. We use higher-dimensional latent representations than in the original disentangled models to preserve signal and motion detail for downstream generation.

Next, *visionECG-Flow* is trained on the latent representations from the pretrained VAEs using the latent rectified flow matching loss defined in Eq. (6),

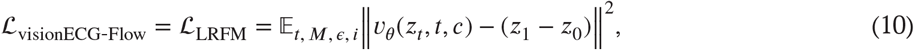

where the flow time *t* ∈ [0, 1]; *M* is a cardiac motion sequence drawn from the data distribution *q*(*M*); ϵ ∼ **N**(**0, 1**) determines the source latent *z*_0_ via Eq. (5); and *i* ∈ {0, 1, …, *T*} selects the target latent *z*_1_: the sequence-level latent *z*_*a*_ for *i* = 0, or the frame-resolved latent *z*_*f*_ for *i* = *f* + 1. *visionECG-Flow* has eight symmetric encoder and decoder residual blocks with a hidden width of 1024 and is optimised using the Adam optimiser (learning rate 5× 10^−5^, 500 epochs). At inference, each flow trajectory is integrated using 100 uniformly spaced explicit Euler steps. For each case, latents are obtained by averaging the terminal latents from 100 independently sampled trajectories^18,36^.

Finally, with *visionECG-Flow* fixed, *visionECG-Motion* is adapted using flow-generated latents so that its training inputs match those encountered at inference. The Transformer and mesh-decoder weights are initialised from the pretrained cardiac motion sequence VAE and the residual modules are newly initialised. All parameters are then jointly optimised on pairs of flow-generated latents and corresponding reference cardiac motion sequences, using the Chamfer-distance and Laplacian terms of Eq. (9),

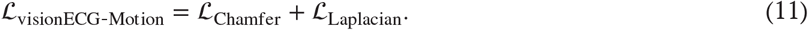

*visionECG-Motion* is trained with the Adam optimiser (learning rate 10^−5^, 100 epochs).

#### ECG signal ablation

To isolate the contribution of subject-specific electrophysiology, we repeated inference with each participant’s ECG latent replaced by the cohort-average ECG latent, leaving all other conditioning unchanged.

#### Method comparison

To our knowledge, no existing model generates a cardiac mesh sequence from an ECG, so we compared *visionECG* against three baselines, each adapted from a related generative method: (i) direct generation from the ECG to the mesh-sequence latent, adapting a convolutional network originally used for ECG latent generation^17^; (ii) a conditional variational autoencoder that decodes meshes from ECG and demographic conditions, adapting a mesh model originally conditioned on demographics alone^18^; and (iii) a contrastive cross-modal generator, adapting a method originally developed for ECG-to-2D four-chamber cardiac MRI synthesis^19^. All baselines used the same inputs, training data and participant splits as *visionECG*.

### Data

Our analysis cohort comprised 71,132 individuals with complete paired data: a resting 12-lead ECG, demographic information, and a time-resolved left-ventricular surface-mesh sequence spanning the cardiac cycle (50 frames; 1,412 vertices and 2,820 faces per frame; Supplementary Fig. 1; Supplementary Table 1). The cohort was partitioned at the participant level into a training set (*n* = 56,906) and a held-out test set (*n* = 14,226), used consistently for all *visionECG* development and downstream clinical analysis, with no participant contributing to more than one split. Within the training set, 2,000 randomly selected cases were reserved for hyperparameter tuning; mesh-generation fidelity was evaluated on 2,000 randomly selected cases from the held-out test set, and the phenotype-agreement analyses used the full test set.

For downstream analyses, disease groups were defined using ICD-10 codes for documented diagnoses of HF, MI, CM, HCM, DCM and hypertension (Supplementary Table 14). MACE was defined as a composite of MI, HF, cardiac arrest, stroke or cardiovascular death (Supplementary Table 14). We additionally defined a control group, free of any cardiac disease, and a healthy group, free of cardiac disease, chronic respiratory disease and metabolic disease (hypercholesterolaemia and diabetes) and with BMI < 30 kg/m^2^(Supplementary Table 15).

For threshold-defined structural abnormality classification, in structural heart disease classification experiments, we included all participants with the required reference phenotypes. For ICD-based disease classification, the analysis included participants with HF, MI, CM, HCM or DCM, together with the control group, which served as a cardiac-disease-free reference for evaluating the discrimination of established cardiac disease. For longitudinal cardiovascular risk stratification, we included all participants with available follow-up data from the groups listed in Supplementary Table 14 to assess incident cardiovascular risk in a broader cohort.

An external dataset from EchoNext-Mini^21,22^of paired echocardiography-derived values and 12-lead ECG signals was used to assess the generalisability of the mesh-based *visionECG*. The validation and test splits of the dataset were used for the external validation. The physical amplitude was restored using the published normalisation constants and resampled from 250 to 500 Hz to match the UK Biobank format, after which the *visionECG* preprocessing pipeline was applied unchanged. An age-matched subcohort of 5,000 participants (median 66 [IQR 60, 72] years; 2,500 per split) was selected to match the UK Biobank (66 [60, 72] years, *P* > 0.05)^40^.

### Structural heart disease classification experiments

We defined structural heart disease in two ways: imaging-based phenotype thresholds, and a documented ICD-coded diagnosis of HF, MI, CM, HCM or DCM. Threshold-defined phenotypes were indexed left ventricular end-diastolic volume (LVEDV) ≥ 62 and ≥ 75 ml/m^2^in women and men, respectively; LVEF ≤ 45% and ≤ 50%; and maximum WT ≥ 13 and ≥ 15 mm^41–43^.

For the threshold tasks, we classified each abnormality directly from the corresponding *visionECG* phenotype (LVEDV, LVEF or maximum WT) using logistic regression, and compared it against three references: (i) a LASSO model of sixteen conventional ECG parameters^44^(Supplementary Fig. 10); (ii) the pretrained EchoNext-Mini model with logistic regression applied to its output probabilities^21,22^; and (iii) EchoNext-Mini with its final layer retrained. The released EchoNext-Mini model outputs only selected binary phenotype probabilities, so it could be applied without retraining only to the LVEF- and WT-based thresholds.^9^

For the ICD-based tasks, we classified each diagnosis from forty-six *visionECG* phenotypes, including twelve global phenotypes, seventeen segmental phenotypes at ED frame and seventeen segmental phenotypes at ES frame, using an XGBoost model^45^(Supplementary Fig. 5), compared against three references: (i) a logistic regression of *visionECG*-derived LVEF and maximum WT; (ii) an XGBoost model of the sixteen ECG parameters; and (iii) EchoNext-Mini with its final layer retrained.

All machine learning models used five-fold cross-validation with class-imbalance weighting. The LASSO penalty was selected by grid search (α ∈ [0, 10]). XGBoost hyperparameters were tuned with Optuna^46^. Each trial sampled maximum depth (1 to 8), learning rate (10^−3^to 3 × 10^−1^), minimum child weight (2 to 200), L1 and L2 regularisation (10^−4^to 10^3^for each) on log-uniform scales, and subsample ratios uniformly (0.3 to 1.0). For EchoNext-Mini retraining, the training set was further split 80%/20% into internal-training and validation subsets, again with class-imbalance weighting.^9^

Discrimination was summarised by the area under the receiver operating characteristic curve (AUC) with 95% confidence intervals; sensitivity and specificity are reported at the Youden-optimal operating point. Models were compared with the two-sided DeLong test^47^.

### Longitudinal cardiovascular risk stratification

We assessed prognostic value for three incident outcomes, HF, MI and MACE, in participants with follow-up records. In each analysis, follow-up began at ECG acquisition and continued until the first event or administrative censoring on 31 October 2022^48^. Participants whose event occurred before ECG acquisition were excluded.

For each outcome, we map the *visionECG* phenotypes or the conventional ECG parameters to a prognostic index by training a XGBoost model with the 5-year event outcome as the output. We compared the two feature groups using Cox PH models. For each index, we report the hazard ratio (HR) per SD with the Wald test and C-index with 95% confidence intervals. To test whether *visionECG* phenotypes add prognostic information beyond conventional ECG parameters, we fitted a nested Cox model containing both prognostic indices from two feature groups and used likelihood-ratio tests to quantify the incremental contribution of each score. Participants were dichotomised at the cohort-median risk score, and Kaplan–Meier curves were compared with the log-rank test^49,50^.

### Statistics

Phenotypes were compared across disease and healthy groups with the Mann–Whitney U test, and *P* values were adjusted for multiple comparisons using the Benjamini–Hochberg false-discovery-rate procedure. An adjusted *P* < 0.05 was considered significant. All analyses used Python (3.10.14) and R (4.5.3).

## Data Availability

All data produced are available online at https://www.ukbiobank.ac.uk/

## Data Availability

All data produced are available online at https://www.ukbiobank.ac.uk/

## Author contributions

Conceptualisation: J.Z., Q.Ma., M.Q., S.K., W.B., D.O.R.; Methodology: J.Z., Q.Ma., M.Q., S.K., Y.M., L.Ch., A.S., M.V., L.Hua., Y.Hu., W.B.; Software: D.S.S., Y.He., M.V.; Formal analysis: J.Z., K.R., M.C., L.Hua., Y.Hu.; Data curation: Q.Me., S.K., W.Z., P.G., N.B., S.T., Y.He., L.Cu., D.S.S.; Writing - original draft: J.Z., L.Hu., Q.Ma., K.R., P.G., L.Hua., M.C., M.V., Y.Hu., D.O.R.; Writing - review & editing: J.Z., S.K., Q.Me., L.Hu., K.R., M.C., N.V., P.G., K.McG., L.Cu., L.Ch., Y.M., A.S., L.Hua., M.Q., Y.Hu., W.B., D.O.R.; Visualisation: J.Z., L.Hu., N.B., K.R., M.Q., Y.Hu., D.O.R.; Supervision: D.O.R., W.B., Y.Hu.; Project administration: D.O.R.; Funding acquisition: D.O.R.

## Data availability

All raw and derived data in this study is available from UK Biobank (http://www.ukbiobank.ac.uk/).

## Code availability

The code used in this analysis is available from GitHub (https://github.com/ImperialCollegeLondon/visionECG) as well as the pipeline for image analysis (https://github.com/ImperialCollegeLondon/DeepMesh).

## Abbreviations

2D: two-dimensional
3D: three-dimensional
4D: four-dimensional
AdaLN: adaptive layer normalisation
ASSD: average symmetric surface distance
AUC: area under the receiver operating characteristic curve
BSA: body surface area
C-index: Harrell’s concordance index
CI: confidence interval
CM: cardiomyopathy
CMR: cardiovascular magnetic resonance
DCM: dilated cardiomyopathy
ECG: electrocardiogram
ED: end-diastolic
EDV: end-diastolic volume
ES: end-systolic
ESV: end-systolic volume
GCS: global circumferential strain
GLS: global longitudinal strain
HCM: hypertrophic cardiomyopathy
HD90: 90^th^percentile Hausdorff distance
HF: heart failure
HR: hazard ratio
ICD: International Classification of Diseases
IQR: interquartile range
KL: Kullback–Leibler
LASSO: least absolute shrinkage and selection operator
LVEDV: left ventricular end-diastolic volume
LVEDVi: left ventricular end-diastolic volume index
LVEF: left ventricular ejection fraction
LVM: left ventricular mass
MACE: major adverse cardiovascular events
MAPSE: mitral annular plane systolic excursion
MI: myocardial infarction
MLP: multilayer perceptron
MSE: mean squared error
PH: proportional hazards
QTc: rate-corrected QT
SD: standard deviation
VAE: variational autoencoder
WT: wall thickness
XGBoost: extreme gradient boosting

