## Supplementary Material for "Reconstructing synthetic hearts from ECG using flow matching"

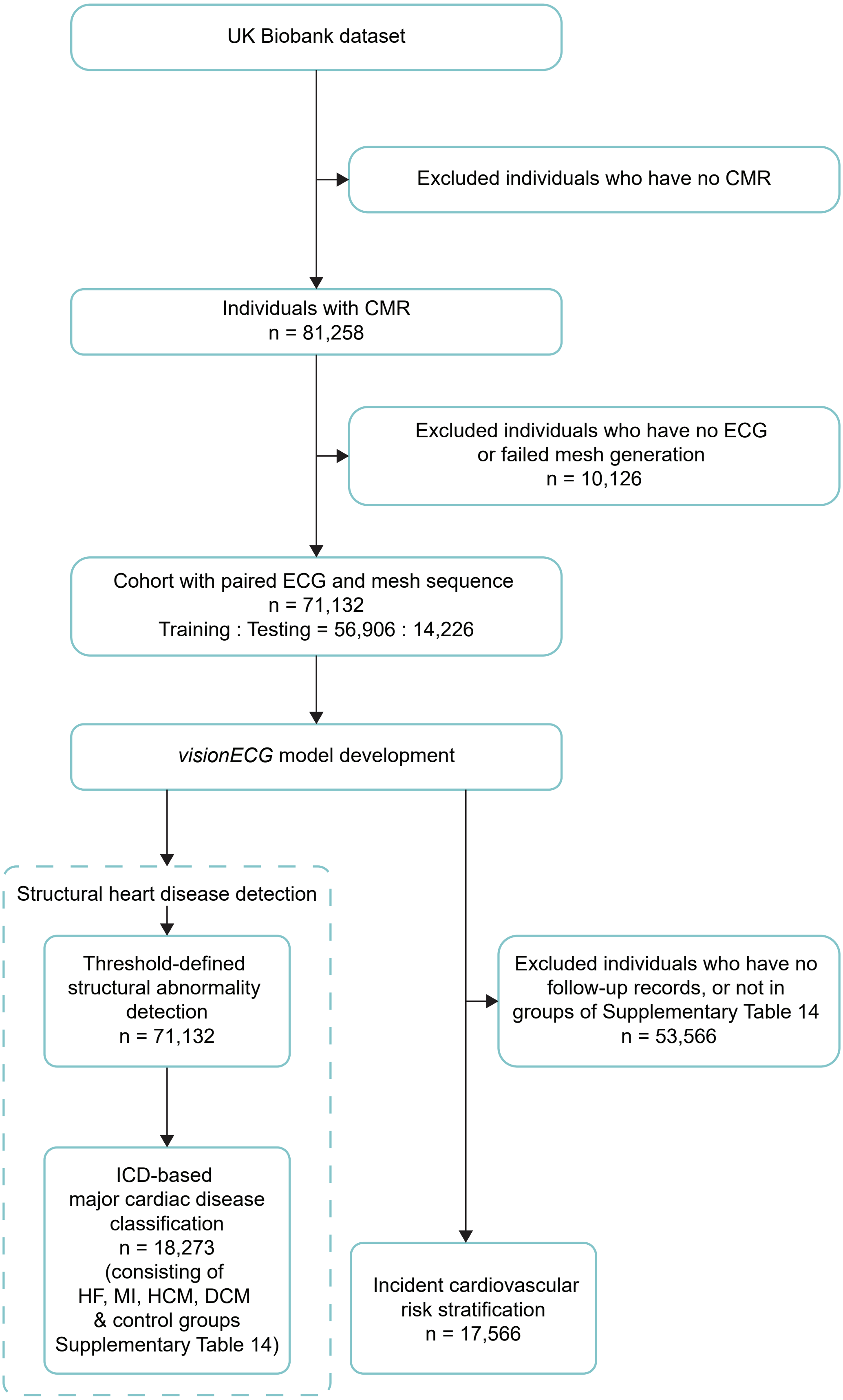


**Supplementary Figure 1: Study flow diagram**. Individual selection to develop *visionECG* model for detection of cardiac structural and functional abnormalities, and its clinical application. CMR, cardiovascular magnetic resonance; ECG, electrocardiogram; HF, heart failure; MI, myocardial infarction; HCM, hypertrophic cardiomyopathy; DCM, dilated cardiomyopathy; MACE, major adverse cardiovascular events; ICD, international classification of diseases.

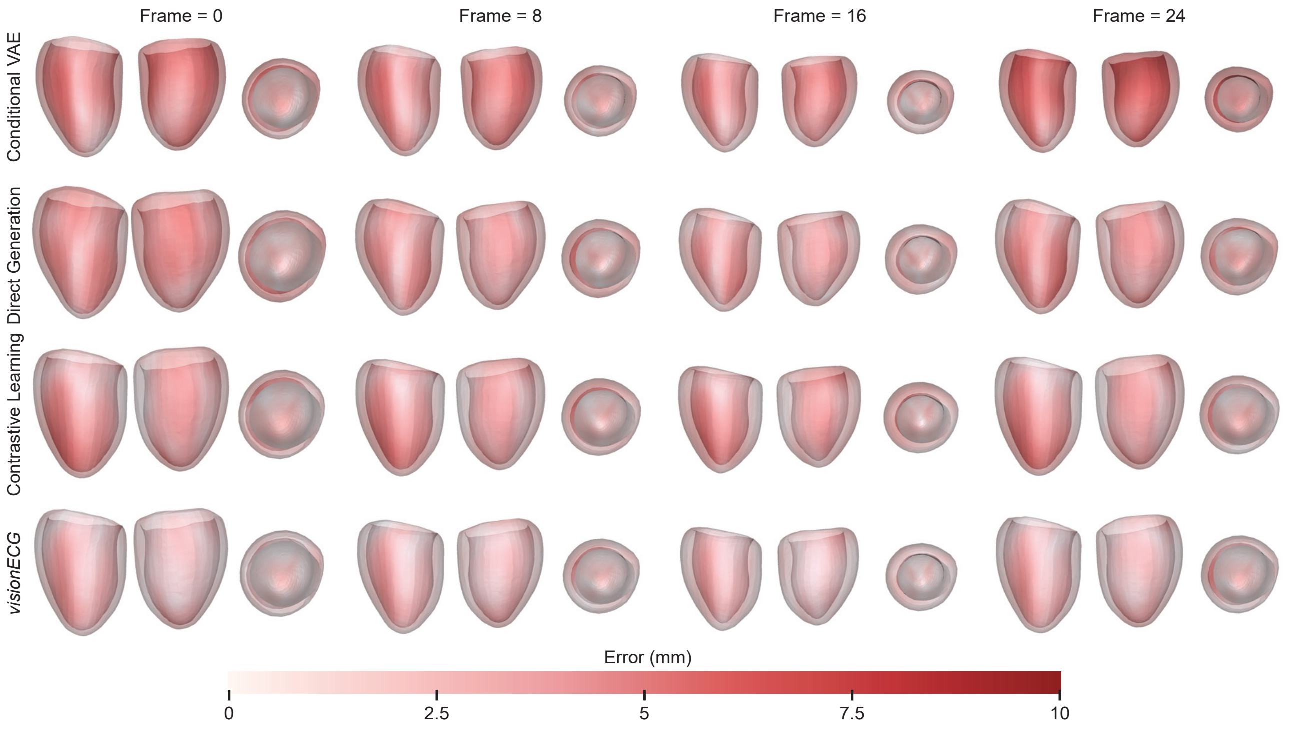


**Supplementary Figure 2: Visualisation of the reconstructed cardiac mesh sequence**. Cardiac meshes generated by *visionECG* and other benchmarks are shown at four time frames. Meshes are coloured by the reconstruction error (in red) with the reference mesh. VAE, variational autoencoder.

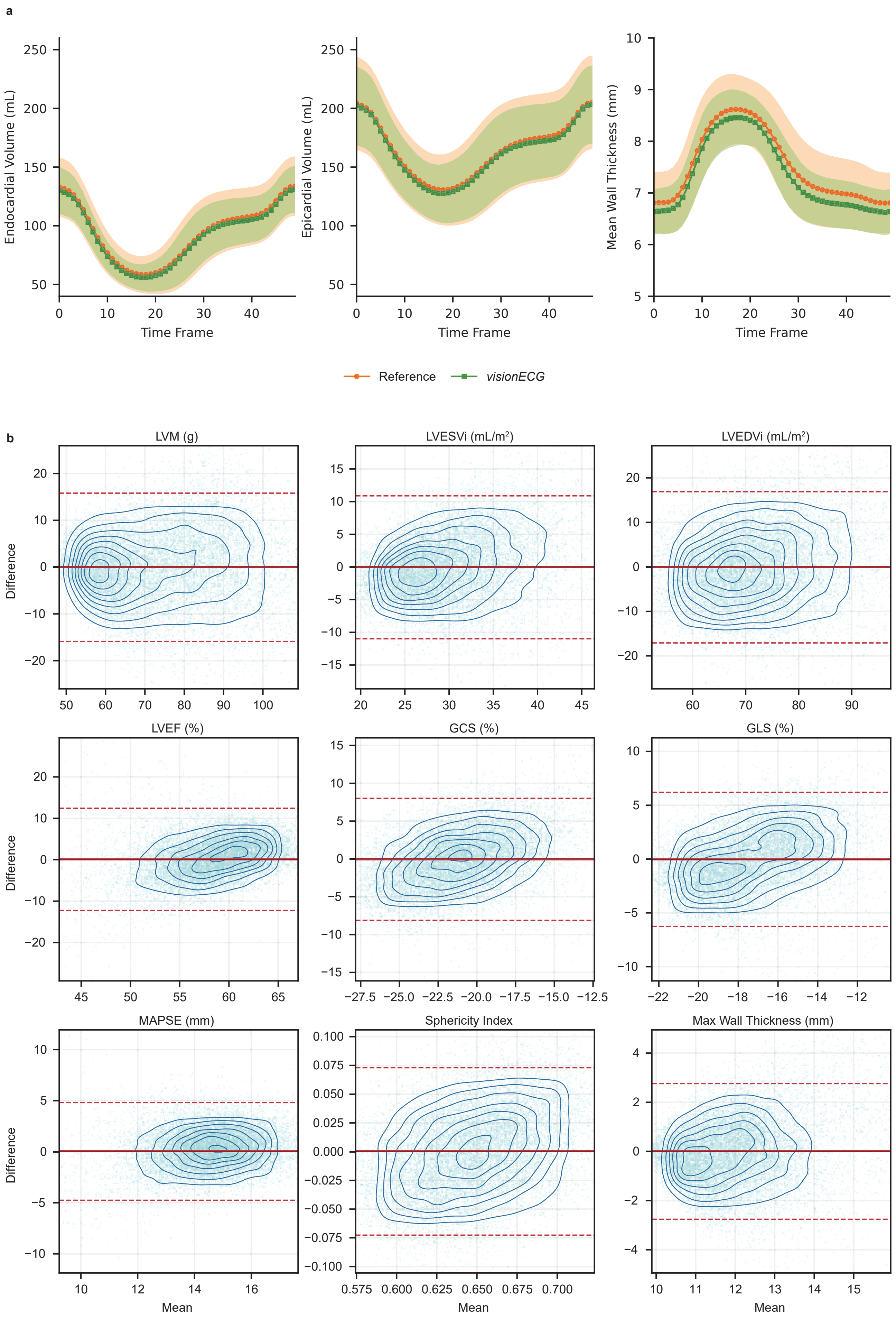


**Supplementary Figure 3: Validation of *visionECG*-derived cardiac phenotypes**. **a**, Volume and wall thickness trajectories are compared between *visionECG* and reference. **b**, Bland-Altman plots assess agreement of *visionECG*-derived measurements with reference. For each plot, solid line, mean bias; dashed lines, 95% limits of agreement. Detailed values are in Supplementary Table 5. LVM, left ventricular mass; LVESVi, left ventricular end-systolic volume index; LVEDVi, left ventricular end-diastolic volume index; LVEF, left ventricular ejection fraction; GCS, global circumferential strain; GLS, global longitudinal strain; MAPSE, mitral annular plane systolic excursion.

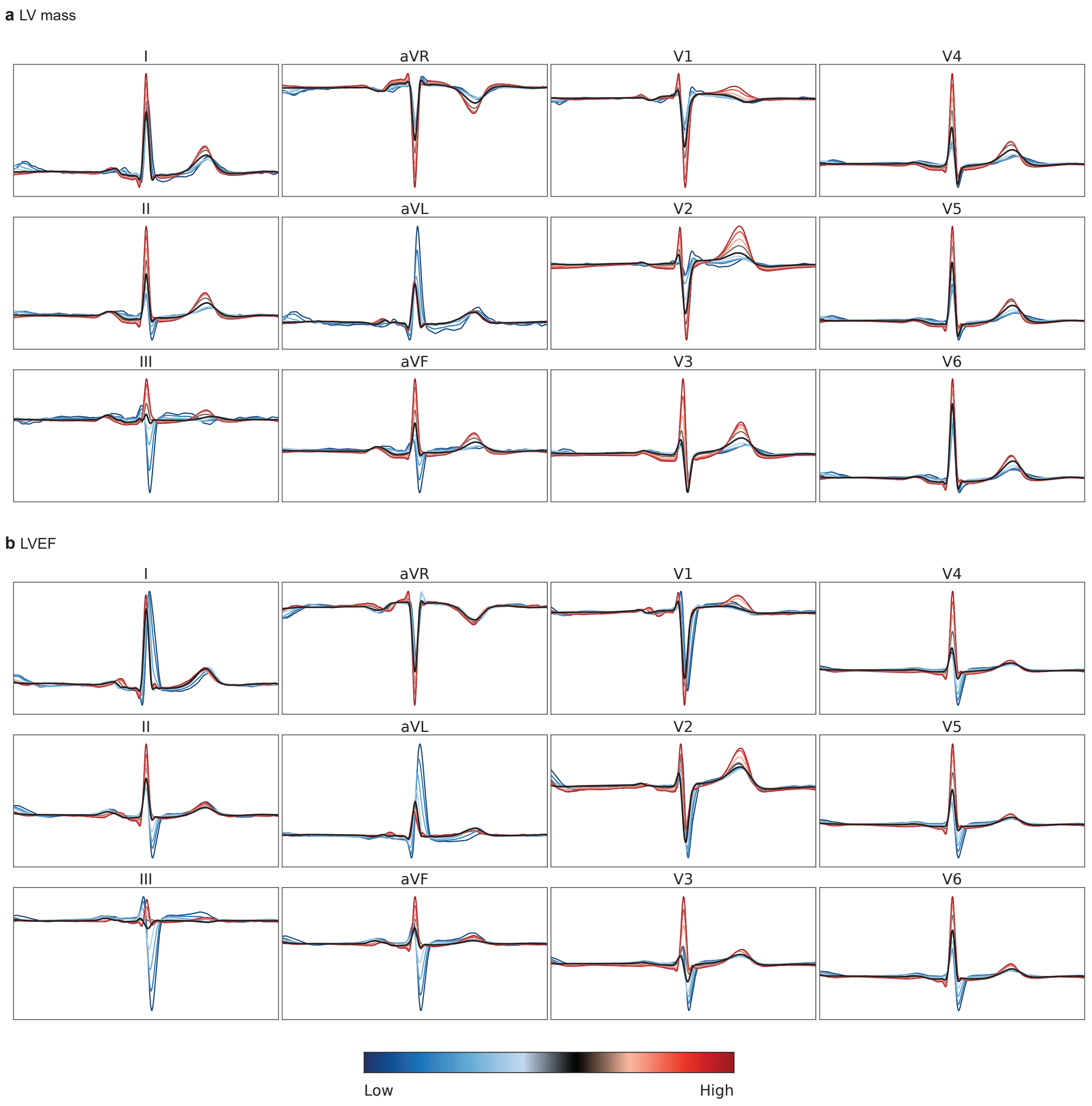


**Supplementary Figure 4: Association between ECG morphologies and *visionECG*-derived cardiac phenotypes**. This example illustrates the association between ECG morphologies and *visionECG*-derived cardiac phenotypes, LV mass (**a**) and LVEF (**b**), with the use of an autoencoder to identify the most important morphological features related to these cardiac phenotypes. LVEF, left ventricular ejection fraction; ECG, electrocardiogram.

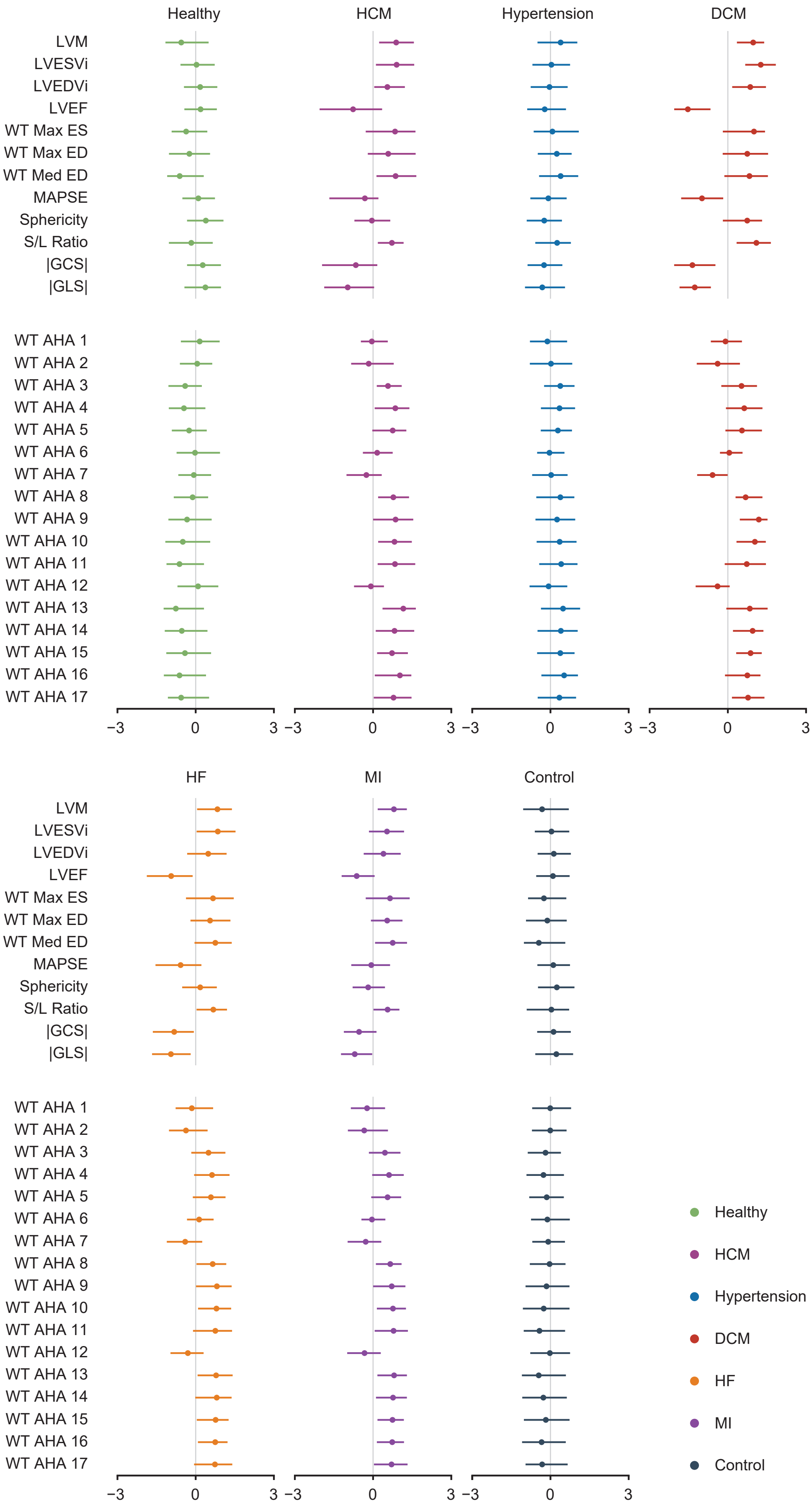


**Supplementary Figure 5: Forest plots of cardiac phenotypes**. Forest plots summarise *visionECG*-derived cardiac phenotype profiles across groups, normalised using the Yeo-Johnson method. Wall thickness measured at segment $N$ at the ED frame is labelled as 'WT AHA $N$'. Points and error bars indicate normalised medians and interquartile ranges. HCM, hypertrophic cardiomyopathy; DCM, dilated cardiomyopathy; HF, heart failure; MI, myocardial infarction; MACE, major adverse cardiovascular events; LVM, left ventricular mass; LVESVi, left ventricular end-systolic volume index; LVEDVi, left ventricular end-diastolic volume index; LVEF, left ventricular ejection fraction; WT, wall thickness; ES, end systole; ED, end diastole; MAPSE, mitral annular plane systolic excursion; S/L, septal-to-lateral ratio; GCS, global circumferential strain; GLS, global longitudinal strain; AHA, American Heart Association.

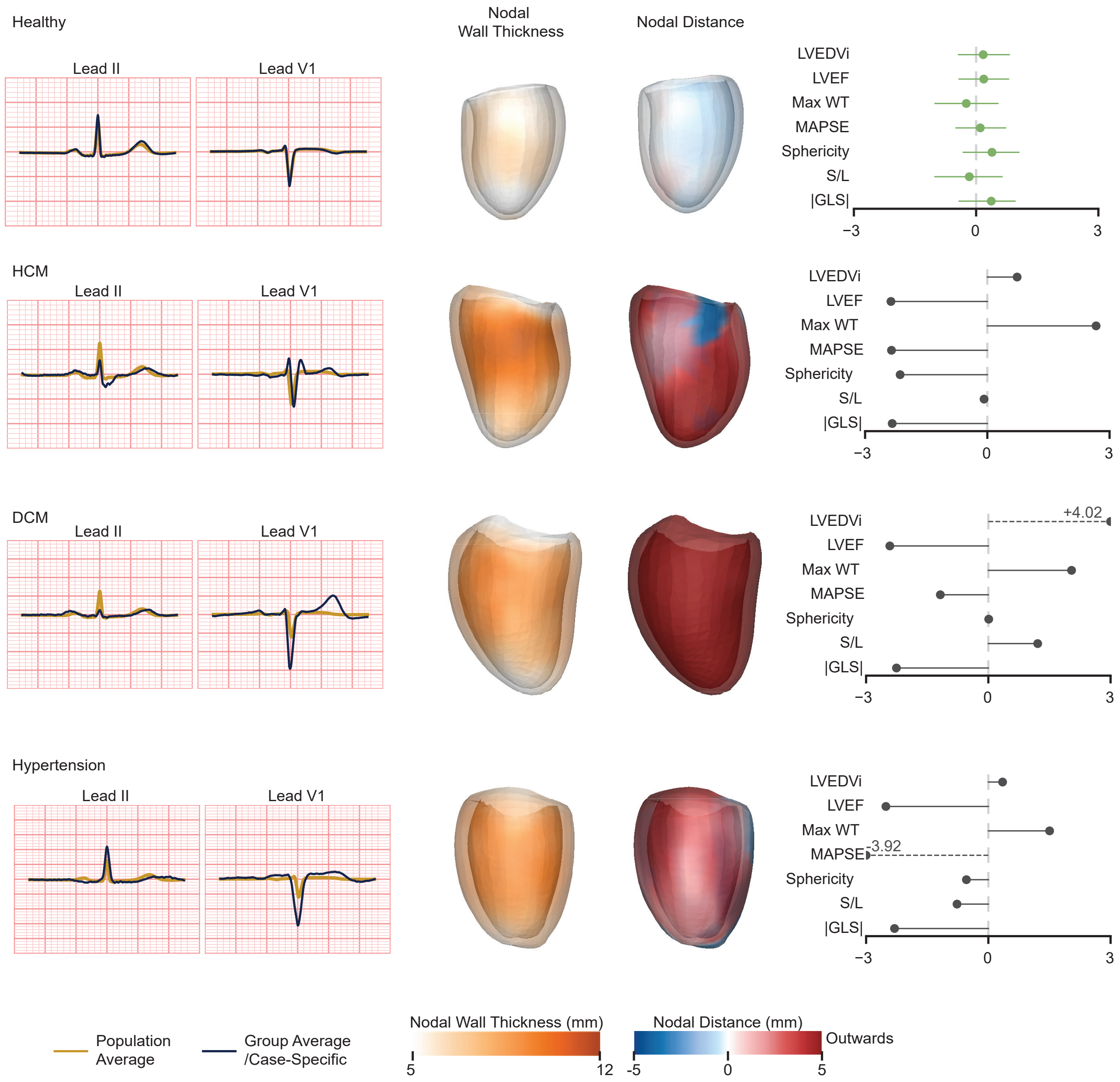


**Supplementary Figure 6: Case-specific cardiac phenotypes**. For representative cases with HCM, DCM and hypertension, left, population-average and case-specific ECG waveforms. Middle, nodal wall thickness and nodal distance mapped onto end-diastolic cardiac meshes. Nodal distance represents the distance of each mesh node relative to the population-average mesh. For the healthy group average meshes in the top row, the averages of nodal wall thickness and nodal distance are displayed. Red and blue colours indicate outward and inward distance, respectively. Right, detailed values of cardiac phenotype measurements. The top row is the healthy group average. HCM, hypertrophic cardiomyopathy; DCM, dilated cardiomyopathy; LVEDVi, left-ventricular end-diastolic volume index; LVEF, left-ventricular ejection fraction; WT, wall thickness; MAPSE, mitral annular plane systolic excursion; S/L, septal-to-lateral ratio; GLS, global longitudinal strain.

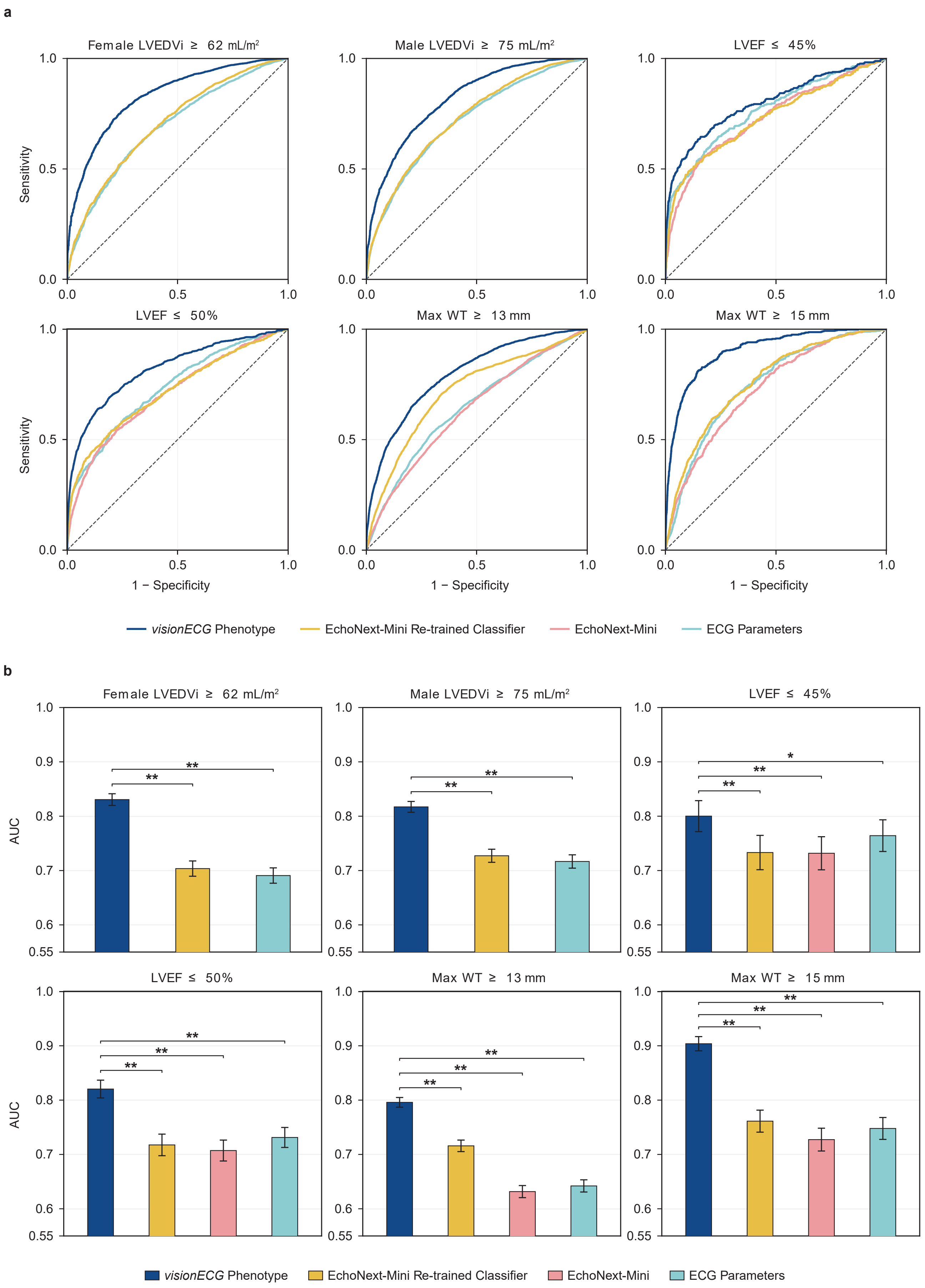


**Supplementary Figure 7: Abnormal phenotype discrimination**. **a**, Receiver operating characteristic curves for threshold-defined abnormalities comparing *visionECG* to other models. **b**, Comparison of AUCs between *visionECG* and other models using the two-sided DeLong test. Specific values refer to Supplementary Table 8. For each plot, ** denotes a notable difference ($P < 0.01$). LVEDVi, left ventricular end-diastolic volume index; LVEF, left ventricular ejection fraction; WT, wall thickness; AUC, area under the receiver operating characteristic curve.

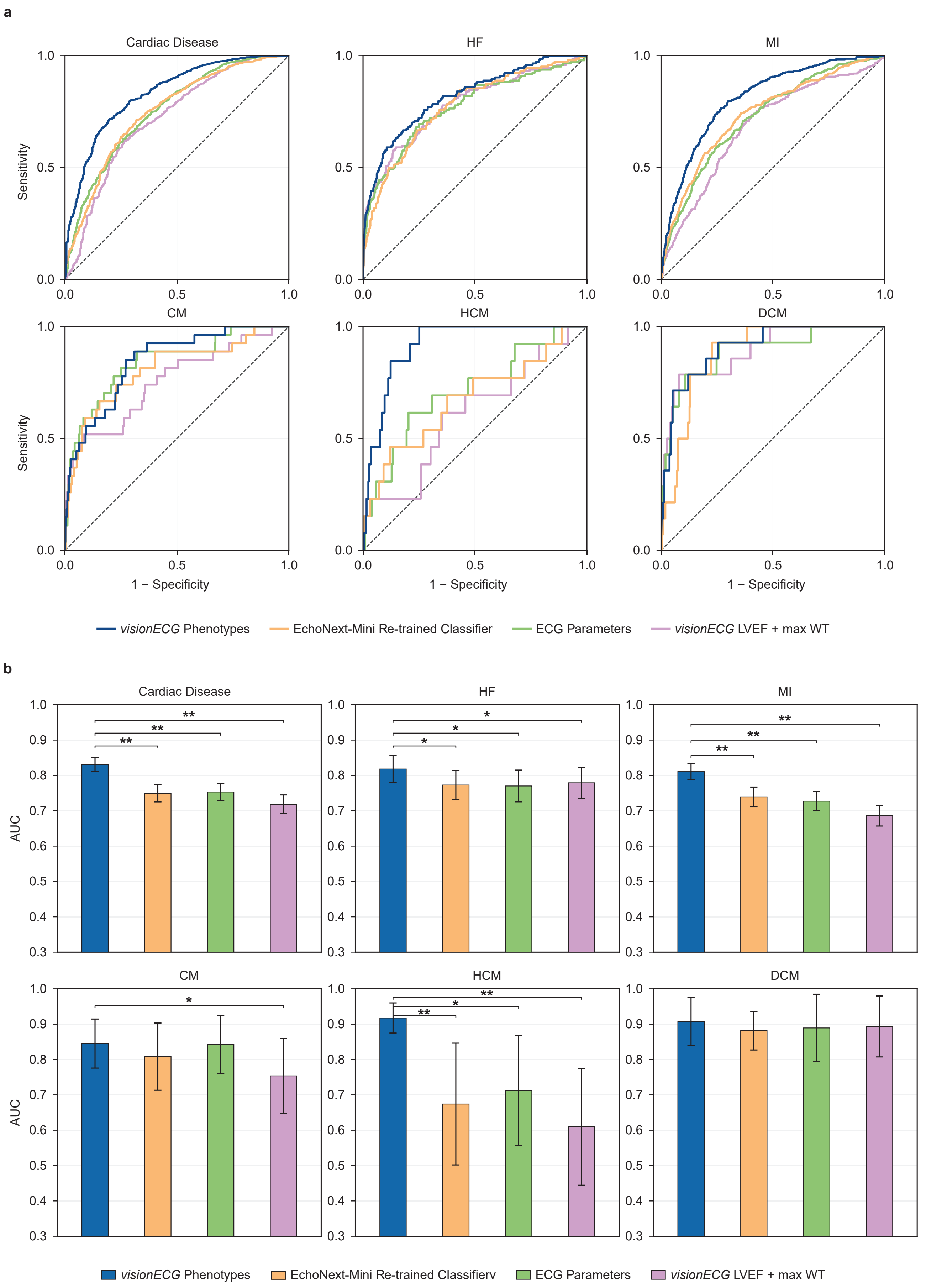


**Supplementary Figure 8: Cardiac disease discrimination**. **a**, Receiver operating characteristic curves for composite and individual cardiac disease comparing *visionECG* to other models. **b**, Comparison of AUCs between *visionECG* and other models using the two-sided DeLong test. Specific values refer to Supplementary Table 10. For each plot, * denotes a notable difference of $P < 0.05$ while ** denotes a notable difference of $P$<0.01. HF, heart failure; MI, myocardial infarction; CM, cardiomyopathy; HCM, hypertrophic cardiomyopathy; DCM, dilated cardiomyopathy; AUC, area under the receiver operating characteristic curve.

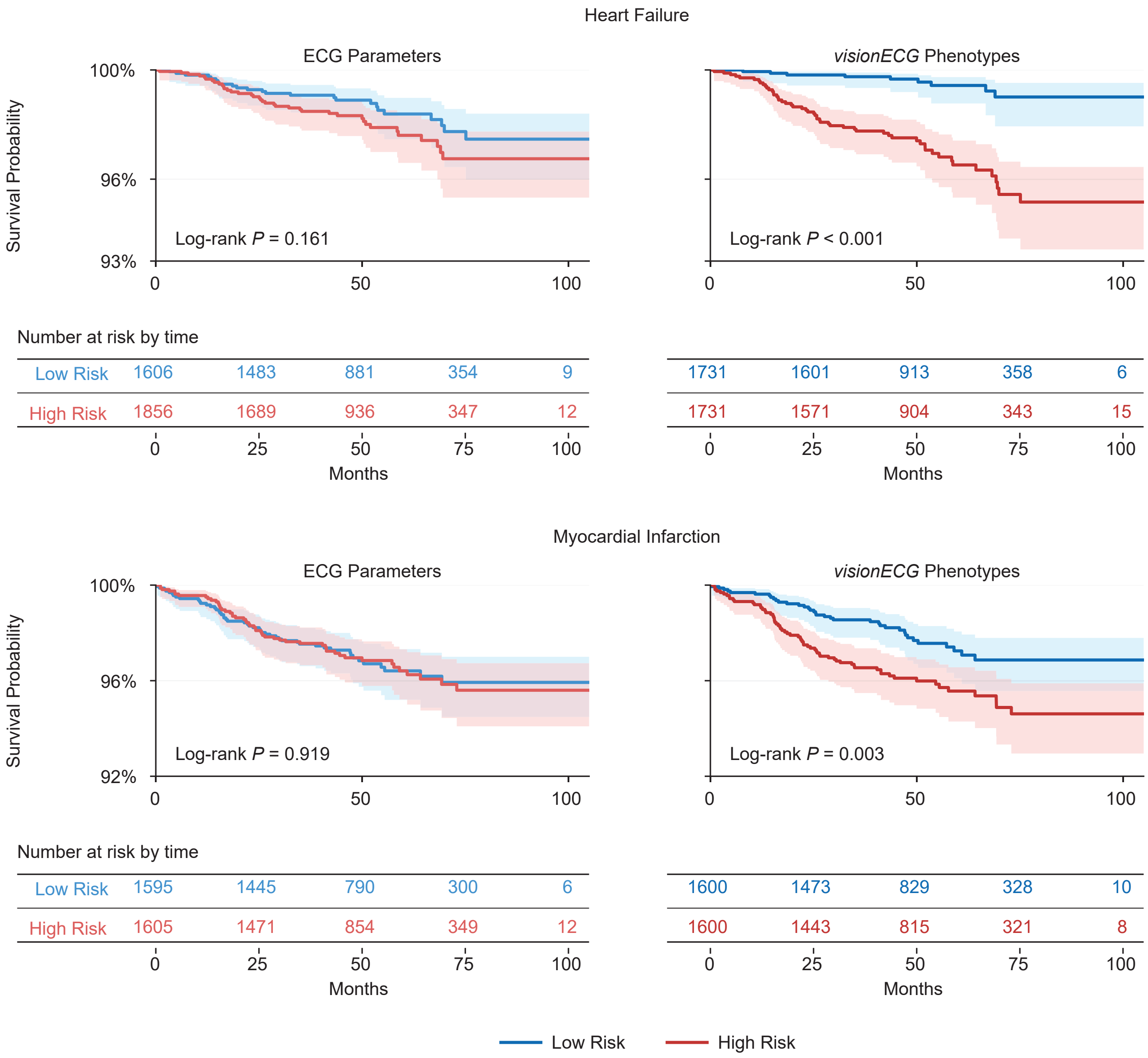


**Supplementary Figure 9: Outcome prediction**. Kaplan-Meier plots for *visionECG* vs. conventional ECG parameters in predicting myocardial infarction and heart failure. For both models, participants were divided into low- and high-risk groups by median risk score. For each plot, the Log-rank test was performed to compare survival between risk groups. HRs with 95% CIs refer to Supplementary Table 12.

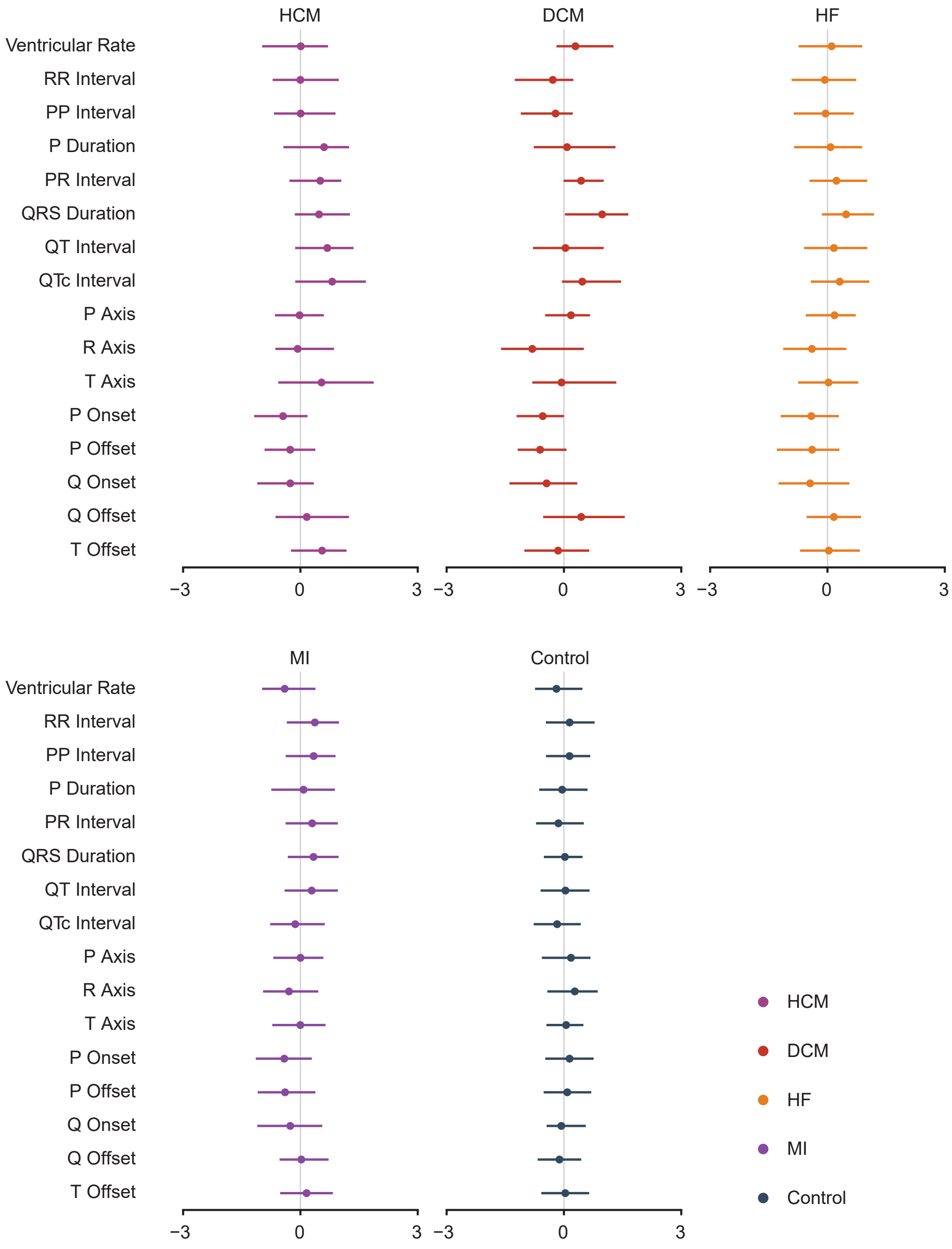


**Supplementary Figure 10: Forest plots of conventional ECG parameters**. Forest plots summarise conventional ECG parameters across groups, normalised using the Yeo-Johnson method. Points and error bars indicate normalised medians and interquartile ranges. HCM, hypertrophic cardiomyopathy; DCM, dilated cardiomyopathy; HF, heart failure; MI, myocardial infarction; MACE, major adverse cardiovascular events.

**Supplementary Table 1: The demographics of participants from the UK Biobank**. Values are presented as n (%) or median [Q1, Q3] unless noted. Control includes individuals without any cardiac disease, and healthy includes individuals without cardiac disease, chronic respiratory disease or metabolic disease, and with BMI $< 30\ \mathrm{kg/m^2}$ (refer to Supplementary Table 15). HF, heart failure; HCM, hypertrophic cardiomyopathy; DCM, dilated cardiomyopathy; MI, myocardial infarction; MRI, magnetic resonance imaging; BSA, body surface area; BMI, body mass index; SBP, systolic blood pressure; DBP, diastolic blood pressure.

| | Overall | HF | HCM | DCM | MI | Hypertension | Control | Healthy |
|---|---|---|---|---|---|---|---|---|
| | n = 71,132 | n = 722 | n = 66 | n = 71 | n = 1,814 | n = 16,706 | n = 15,953 | n = 4,449 |
| Sex Males (n, %) | 32866 (46.2%) | 490 (67.9%) | 46 (69.7%) | 52 (73.2%) | 1411 (77.8%) | 9179 (54.9%) | 6428 (40.3%) | 1603 (36.0%) |
| Ethnicity White (n, %) | 68512 (96.3%) | 701 (97.1%) | 65 (98.5%) | 70 (98.6%) | 1747 (96.3%) | 16014 (95.9%) | 15515 (97.3%) | 4335 (97.4%) |
| Age at time of CMR (years) | 66.00 [60.00, 72.00] | 72.00 [67.00, 76.00] | 67.00 [62.00, 72.75] | 71.00 [66.00, 75.00] | 71.00 [65.00, 75.00] | 69.00 [63.00, 74.00] | 63.00 [56.00, 68.00] | 61.00 [56.00, 67.00] |
| BSA ($m^2$) | 1.86 [1.71, 2.01] | 1.95 [1.80, 2.11] | 1.93 [1.78, 2.06] | 1.98 [1.85, 2.11] | 1.94 [1.82, 2.08] | 1.92 [1.76, 2.07] | 1.81 [1.68, 1.96] | 1.77 [1.64, 1.91] |
| BMI ($kg/m^2$) | 25.90 [23.50, 28.70] | 26.90 [24.50, 30.30] | 26.95 [24.50, 30.20] | 27.60 [25.10, 30.10] | 27.00 [24.60, 29.80] | 27.30 [24.70, 30.30] | 24.90 [22.80, 27.30] | 24.10 [22.20, 26.20] |
| SBP (mmHg) | 140.50 [128.00, 154.00] | 140.50 [127.50, 155.00] | 142.75 [129.25, 159.62] | 134.00 [122.00, 148.50] | 142.00 [129.00, 155.00] | 148.00 [136.50, 160.50] | 133.50 [122.50, 145.50] | 132.00 [121.00, 144.50] |
| DBP (mmHg) | 78.50 [72.00, 85.50] | 76.50 [69.50, 85.50] | 77.50 [71.62, 87.00] | 75.50 [70.75, 85.50] | 76.50 [70.00, 84.00] | 81.50 [74.50, 88.00] | 76.50 [70.50, 83.50] | 76.00 [70.00, 83.00] |

**Supplementary Table 2: Comparison of cardiac volume and wall thickness errors**. Volume errors are reported as absolute difference (mL) and absolute relative difference (%). Wall thickness errors are reported as absolute difference (mm) and absolute relative difference (%). All values are median[IQR]; lower is better. Paired Wilcoxon signed-rank $P$-values vs. *visionECG* are shown next to each metric. Underlined metric values mark the best (minimum) within each section. $P$-values are bold when $P < 0.05$.

| Methods | Endocardial Volume Absolute Difference (mL)↓ | | Endocardial Volume Absolute Relative Difference (%)↓ | | Wall Thickness Absolute Difference (mm)↓ | | Wall Thickness Absolute Relative Difference (%)↓ | |
|---|---|---|---|---|---|---|---|---|
| | Value | $P$ | Value | $P$ | Value | $P$ | Value | $P$ |
| **Mean across sequence** | | | | | | | | |
| Direct Generation | $13.865_{[8.169,21.461]}$ | $\mathbf{< 10^{-16}}$ | $13.426_{[8.059,22.358]}$ | $\mathbf{< 10^{-16}}$ | $0.840_{[0.703,1.033]}$ | $\mathbf{< 10^{-16}}$ | $11.871_{[10.170,14.227]}$ | $\mathbf{< 10^{-16}}$ |
| Conditioned VAE | $10.532_{[5.976,18.381]}$ | $\mathbf{< 10^{-16}}$ | $11.522_{[6.716,18.544]}$ | $\mathbf{< 10^{-16}}$ | $0.758_{[0.641,0.923]}$ | $\mathbf{< 10^{-16}}$ | $10.763_{[9.274,12.830]}$ | $\mathbf{< 10^{-16}}$ |
| Contrastive Learning | $9.742_{[5.345,17.147]}$ | $\mathbf{< 10^{-16}}$ | $10.542_{[6.241,17.446]}$ | $\mathbf{< 10^{-16}}$ | $0.724_{[0.626,0.867]}$ | $\mathbf{< 10^{-16}}$ | $10.262_{[8.912,12.086]}$ | $\mathbf{< 10^{-16}}$ |
| *visionECG* | $\underline{7.839_{[4.443,13.398]}}$ | — | $\underline{8.341_{[5.077,13.504]}}$ | — | $\underline{0.679_{[0.581,0.804]}}$ | — | $\underline{9.535_{[8.224,11.143]}}$ | — |
| **ED frame** | | | | | | | | |
| Direct Generation | $17.054_{[8.166,26.172]}$ | $\mathbf{< 10^{-16}}$ | $12.308_{[5.805,21.777]}$ | $\mathbf{< 10^{-16}}$ | $0.814_{[0.655,1.038]}$ | $\mathbf{< 10^{-16}}$ | $12.587_{[10.191,15.808]}$ | $\mathbf{< 10^{-16}}$ |
| Conditioned VAE | $11.687_{[5.610,20.793]}$ | $\mathbf{< 10^{-16}}$ | $9.155_{[4.331,15.641]}$ | $\mathbf{< 10^{-16}}$ | $0.705_{[0.582,0.877]}$ | $\mathbf{< 10^{-16}}$ | $11.117_{[9.266,13.532]}$ | $\mathbf{< 10^{-16}}$ |
| Contrastive Learning | $11.509_{[5.140,19.943]}$ | $\mathbf{< 10^{-16}}$ | $8.868_{[4.038,14.653]}$ | $\mathbf{< 10^{-16}}$ | $0.678_{[0.564,0.839]}$ | $\mathbf{< 10^{-16}}$ | $10.695_{[8.948,12.921]}$ | $\mathbf{< 10^{-16}}$ |
| *visionECG* | $\underline{9.125_{[4.219,15.598]}}$ | — | $\underline{6.906_{[3.298,11.648]}}$ | — | $\underline{0.630_{[0.526,0.760]}}$ | — | $\underline{9.852_{[8.227,11.687]}}$ | — |
| **ES frame** | | | | | | | | |
| Direct Generation | $7.176_{[3.406,11.412]}$ | $\mathbf{< 10^{-16}}$ | $13.343_{[6.263,21.699]}$ | $\mathbf{< 10^{-16}}$ | $0.786_{[0.658,0.972]}$ | $\mathbf{< 10^{-16}}$ | $9.537_{[8.076,11.472]}$ | $\mathbf{< 10^{-16}}$ |
| Conditioned VAE | $6.462_{[3.083,10.928]}$ | $\mathbf{< 10^{-16}}$ | $12.176_{[5.565,21.105]}$ | $\mathbf{< 10^{-16}}$ | $0.750_{[0.617,0.914]}$ | $\mathbf{< 10^{-16}}$ | $9.071_{[7.626,10.861]}$ | $\mathbf{< 10^{-16}}$ |
| Contrastive Learning | $6.193_{[3.094,10.971]}$ | $\mathbf{< 10^{-16}}$ | $11.355_{[5.671,19.760]}$ | $\mathbf{< 10^{-16}}$ | $0.728_{[0.615,0.881]}$ | $\mathbf{< 10^{-16}}$ | $8.792_{[7.425,10.431]}$ | $\mathbf{< 10^{-16}}$ |
| *visionECG* | $\underline{4.880_{[2.328,8.604]}}$ | — | $\underline{8.923_{[4.442,15.661]}}$ | — | $\underline{0.688_{[0.580,0.836]}}$ | — | $\underline{8.272_{[7.053,9.898]}}$ | — |

**Supplementary Table 3: Comparison of cardiac mesh reconstruction accuracy (HD90 and ASSD)**. The Hausdorff distance, 90th percentile (HD90) and average symmetric surface distance (ASSD) are reported for the whole sequence and at the end-diastolic (ED) and end-systolic (ES) frames. Values are reported as median[IQR]; lower is better. Paired Wilcoxon signed-rank $P$-values vs. *visionECG* are shown next to each metric. Underlined metric values mark the best (minimum) within each section. $P$-values are bold when $P < 0.05$. Reduced view: LVendo and LVepi only.

| Methods | HD90 (*mm*)↓ | | | | ASSD (*mm*)↓ | | | |
|---|---|---|---|---|---|---|---|---|
| | LVendo | | LVepi | | LVendo | | LVepi | |
| | Value | $P$ | Value | $P$ | Value | $P$ | Value | $P$ |
| **Mean across sequence** | | | | | | | | |
| Direct Generation | $4.050_{[3.530,4.692]}$ | $\mathbf{< 10^{-16}}$ | $4.259_{[3.738,4.998]}$ | $\mathbf{< 10^{-16}}$ | $2.479_{[2.214,2.807]}$ | $\mathbf{< 10^{-16}}$ | $2.640_{[2.374,2.986]}$ | $\mathbf{< 10^{-16}}$ |
| Conditioned VAE | $3.632_{[3.220,4.265]}$ | $\mathbf{< 10^{-16}}$ | $3.769_{[3.345,4.401]}$ | $\mathbf{< 10^{-16}}$ | $2.293_{[2.063,2.638]}$ | $\mathbf{< 10^{-16}}$ | $2.395_{[2.149,2.741]}$ | $\mathbf{< 10^{-16}}$ |
| Contrastive Learning | $3.476_{[3.100,4.047]}$ | $\mathbf{< 10^{-16}}$ | $3.679_{[3.267,4.252]}$ | $\mathbf{< 10^{-16}}$ | $2.199_{[1.988,2.511]}$ | $\mathbf{< 10^{-16}}$ | $2.360_{[2.118,2.693]}$ | $\mathbf{< 10^{-16}}$ |
| *visionECG* | $\underline{3.211_{[2.909,3.618]}}$ | — | $\underline{3.365_{[3.024,3.797]}}$ | — | $\underline{2.061_{[1.878,2.282]}}$ | — | $\underline{2.185_{[1.971,2.431]}}$ | — |
| **ED frame** | | | | | | | | |
| Direct Generation | $3.795_{[3.300,4.603]}$ | $\mathbf{< 10^{-16}}$ | $4.133_{[3.556,4.994]}$ | $\mathbf{< 10^{-16}}$ | $2.428_{[2.157,2.798]}$ | $\mathbf{< 10^{-16}}$ | $2.640_{[2.326,3.025]}$ | $\mathbf{< 10^{-16}}$ |
| Conditioned VAE | $3.272_{[2.978,3.805]}$ | $\mathbf{< 10^{-16}}$ | $3.539_{[3.158,4.133]}$ | $\mathbf{< 10^{-16}}$ | $2.162_{[1.944,2.452]}$ | $\mathbf{< 10^{-16}}$ | $2.311_{[2.054,2.655]}$ | $\mathbf{< 10^{-16}}$ |
| Contrastive Learning | $3.260_{[2.949,3.749]}$ | $\mathbf{< 10^{-16}}$ | $3.539_{[3.110,4.045]}$ | $\mathbf{< 10^{-16}}$ | $2.127_{[1.921,2.420]}$ | $\mathbf{< 10^{-16}}$ | $2.312_{[2.058,2.622]}$ | $\mathbf{< 10^{-16}}$ |
| *visionECG* | $\underline{3.051_{[2.802,3.373]}}$ | — | $\underline{3.245_{[2.916,3.626]}}$ | — | $\underline{1.992_{[1.832,2.193]}}$ | — | $\underline{2.131_{[1.928,2.382]}}$ | — |
| **ES frame** | | | | | | | | |
| Direct Generation | $3.668_{[3.132,4.434]}$ | $\mathbf{< 10^{-16}}$ | $4.076_{[3.487,4.866]}$ | $\mathbf{< 10^{-16}}$ | $2.204_{[1.936,2.559]}$ | $\mathbf{< 10^{-16}}$ | $2.522_{[2.230,2.912]}$ | $\mathbf{< 10^{-16}}$ |
| Conditioned VAE | $3.444_{[2.953,4.082]}$ | $\mathbf{< 10^{-16}}$ | $3.761_{[3.280,4.450]}$ | $\mathbf{< 10^{-16}}$ | $2.119_{[1.856,2.443]}$ | $\mathbf{< 10^{-16}}$ | $2.374_{[2.110,2.738]}$ | $\mathbf{< 10^{-16}}$ |
| Contrastive Learning | $3.383_{[2.897,4.059]}$ | $\mathbf{< 10^{-16}}$ | $3.723_{[3.244,4.438]}$ | $\mathbf{< 10^{-16}}$ | $2.082_{[1.832,2.440]}$ | $\mathbf{< 10^{-16}}$ | $2.374_{[2.096,2.739]}$ | $\mathbf{< 10^{-16}}$ |
| *visionECG* | $\underline{3.099_{[2.722,3.674]}}$ | — | $\underline{3.427_{[3.004,3.958]}}$ | — | $\underline{1.952_{[1.725,2.244]}}$ | — | $\underline{2.190_{[1.948,2.490]}}$ | — |

**Supplementary Table 4: ECG ablation of *visionECG* on volume errors.** Volume errors are reported as absolute difference (mL) and absolute relative difference (%). Wall thickness errors are reported as absolute difference (mm) and absolute relative difference (%). All values are median[IQR]; lower is better. Paired Wilcoxon signed-rank $P$-values for *visionECG* vs. ECG ablation are shown for each disease row. Underlined metric values mark the better arm within each disease row. $P$-values are bold when $P < 0.05$.

| Disease | Endocardial Volume Absolute Difference (mL)↓ | | | Endocardial Volume Absolute Relative Difference (%)↓ | | |
|---|---|---|---|---|---|---|
| | ECG Ablation | *visionECG* | $P$ | ECG Ablation | *visionECG* | $P$ |
| **Whole Sequence** | | | | | | |
| Healthy | $9.646_{[5.729,16.118]}$ | $\underline{7.080}_{[4.056,12.435]}$ | $\mathbf{< 10^{-16}}$ | $10.328_{[6.405,16.546]}$ | $\underline{7.638}_{[4.542,12.321]}$ | $\mathbf{< 10^{-16}}$ |
| DCM | $24.594_{[14.293,39.161]}$ | $\underline{14.338}_{[10.044,24.197]}$ | $\mathbf{5.1 \times 10^{-9}}$ | $20.291_{[13.504,28.544]}$ | $\underline{12.592}_{[8.725,17.934]}$ | $\mathbf{9.1 \times 10^{-9}}$ |
| HCM | $15.960_{[8.218,23.504]}$ | $\underline{9.608}_{[6.235,16.382]}$ | **0.0042** | $14.859_{[8.647,22.971]}$ | $\underline{10.249}_{[5.209,16.861]}$ | **0.0099** |
| HF | $17.858_{[9.376,34.441]}$ | $\underline{11.646}_{[6.673,20.842]}$ | $\mathbf{< 10^{-16}}$ | $16.971_{[9.814,27.338]}$ | $\underline{10.654}_{[6.393,17.249]}$ | $\mathbf{< 10^{-16}}$ |
| **ED Frame** | | | | | | |
| Healthy | $11.197_{[5.534,18.802]}$ | $\underline{8.159}_{[3.687,14.347]}$ | $\mathbf{< 10^{-16}}$ | $8.881_{[4.302,14.661]}$ | $\underline{6.408}_{[2.974,11.012]}$ | $\mathbf{< 10^{-16}}$ |
| DCM | $20.995_{[8.929,40.740]}$ | $\underline{14.976}_{[7.685,25.672]}$ | $\mathbf{4.0 \times 10^{-4}}$ | $13.960_{[5.933,23.886]}$ | $\underline{10.287}_{[5.291,15.771]}$ | $\mathbf{6.9 \times 10^{-4}}$ |
| HCM | $16.775_{[7.002,29.928]}$ | $\underline{11.229}_{[5.339,22.365]}$ | **0.0151** | $11.424_{[4.992,20.317]}$ | $\underline{7.751}_{[3.488,16.478]}$ | **0.0311** |
| HF | $17.806_{[8.535,33.703]}$ | $\underline{12.489}_{[5.518,22.475]}$ | $\mathbf{< 10^{-16}}$ | $12.274_{[6.347,21.301]}$ | $\underline{8.843}_{[3.923,14.536]}$ | $\mathbf{< 10^{-16}}$ |
| **ES Frame** | | | | | | |
| Healthy | $6.133_{[2.873,10.364]}$ | $\underline{4.427}_{[2.073,7.746]}$ | $\mathbf{< 10^{-16}}$ | $11.541_{[5.304,20.226]}$ | $\underline{8.435}_{[3.971,14.951]}$ | $\mathbf{< 10^{-16}}$ |
| DCM | $17.618_{[8.584,36.480]}$ | $\underline{9.867}_{[6.471,18.288]}$ | $\mathbf{2.0 \times 10^{-8}}$ | $23.054_{[13.339,35.477]}$ | $\underline{13.535}_{[8.656,23.412]}$ | $\mathbf{9.3 \times 10^{-8}}$ |
| HCM | $10.217_{[5.179,19.983]}$ | $\underline{7.230}_{[3.932,11.588]}$ | **0.0046** | $17.423_{[8.033,26.172]}$ | $\underline{12.105}_{[5.769,16.331]}$ | **0.0070** |
| HF | $11.222_{[5.401,24.514]}$ | $\underline{7.640}_{[3.142,15.471]}$ | $\mathbf{< 10^{-16}}$ | $17.831_{[9.227,31.610]}$ | $\underline{11.851}_{[5.026,20.496]}$ | $\mathbf{< 10^{-16}}$ |

**Supplementary Table 5: Bland-Altman analysis results**. Bland-Altman analysis assessed agreement of *visionECG*-derived measurements with reference and detailed values are reported. Bias, mean difference between *visionECG*-derived measurements and reference; (95%) limits of agreement, bias±1.96 s.d. of the differences. GCS, global circumferential strain; GLS, global longitudinal strain; MAPSE, mitral annular plane systolic excursion; LVEDVi, left ventricular end-diastolic volume index; LVM, left ventricular mass; LVESVi, left ventricular end-systolic volume index; LVEF, left ventricular ejection fraction.

| Measurement | Bias | Limits of agreement |
|---|---|---|
| GCS (%) | -0.057 | [-8.122, 8.007] |
| GLS (%) | -0.028 | [-6.257, 6.201] |
| MAPSE (mm) | 0.031 | [-4.752, 4.814] |
| LVEDVi (ml/m$^2$) | -0.097 | [-17.086, 16.893] |
| Max wall thickness (mm) | $6.7 \times 10^{-4}$ | [-2.759, 2.760] |
| LVM (g) | -0.043 | [-15.892, 15.806] |
| LVEF (%) | 0.087 | [-12.255, 12.430] |
| LVESVi (ml/m$^2$) | -0.052 | [-10.992, 10.888] |
| Sphericity index | $1.8 \times 10^{-4}$ | [-0.073, 0.073] |

**Supplementary Table 6: Comparison of *visionECG*-derived phenotypes between healthy and each disease group**. Benjamini-Hochberg corrected Mann-Whitney $P$-values are reported for each comparison. $P$-values are bold when $P < 0.05$. DCM, dilated cardiomyopathy; HCM, hypertrophic cardiomyopathy; HF, heart failure; LVM, left ventricular mass; LVEDV, left ventricular end-diastolic volume; LVESV, left ventricular end-systolic volume; LVEDVi, left ventricular end-diastolic volume index; LVESVi, left ventricular end-systolic volume index; LVEF, left ventricular ejection fraction; WT, wall thickness; ES, end systole; ED, end diastole; MAPSE, mitral annular plane systolic excursion; S/L, septal-to-lateral ratio; GCS, global circumferential strain; GLS, global longitudinal strain.

| Feature | Healthy & DCM | Healthy & HCM | Healthy & Hypertension | Healthy & HF |
|---|---|---|---|---|
| LVM | $\mathbf{< 10^{-16}}$ | $\mathbf{2.2 \times 10^{-16}}$ | $\mathbf{< 10^{-16}}$ | $\mathbf{< 10^{-16}}$ |
| LVEDV | $\mathbf{6.0 \times 10^{-15}}$ | $\mathbf{1.8 \times 10^{-9}}$ | $\mathbf{< 10^{-16}}$ | $\mathbf{< 10^{-16}}$ |
| LVESV | $\mathbf{< 10^{-16}}$ | $\mathbf{2.2 \times 10^{-12}}$ | $\mathbf{< 10^{-16}}$ | $\mathbf{< 10^{-16}}$ |
| LVEDVi | **0.0022** | **0.0019** | $\mathbf{< 10^{-16}}$ | $\mathbf{5.3 \times 10^{-9}}$ |
| LVESVi | $\mathbf{< 10^{-16}}$ | $\mathbf{5.3 \times 10^{-9}}$ | 0.1793 | $\mathbf{< 10^{-16}}$ |
| LVEF | $\mathbf{< 10^{-16}}$ | $\mathbf{1.0 \times 10^{-11}}$ | $\mathbf{< 10^{-16}}$ | $\mathbf{< 10^{-16}}$ |
| WT Max ES | $\mathbf{4.5 \times 10^{-9}}$ | $\mathbf{1.1 \times 10^{-10}}$ | $\mathbf{< 10^{-16}}$ | $\mathbf{< 10^{-16}}$ |
| WT Max ED | $\mathbf{2.1 \times 10^{-12}}$ | $\mathbf{2.5 \times 10^{-10}}$ | $\mathbf{< 10^{-16}}$ | $\mathbf{< 10^{-16}}$ |
| WT Med ED | $\mathbf{< 10^{-16}}$ | $\mathbf{< 10^{-16}}$ | $\mathbf{< 10^{-16}}$ | $\mathbf{< 10^{-16}}$ |
| MAPSE | $\mathbf{< 10^{-16}}$ | $\mathbf{8.8 \times 10^{-8}}$ | $\mathbf{< 10^{-16}}$ | $\mathbf{< 10^{-16}}$ |
| Sphericity | 0.2104 | **0.0028** | $\mathbf{< 10^{-16}}$ | $\mathbf{5.5 \times 10^{-6}}$ |
| S/L | $\mathbf{1.9 \times 10^{-15}}$ | $\mathbf{1.6 \times 10^{-8}}$ | $\mathbf{< 10^{-16}}$ | $\mathbf{< 10^{-16}}$ |
| GCS | $\mathbf{< 10^{-16}}$ | $\mathbf{1.2 \times 10^{-13}}$ | $\mathbf{< 10^{-16}}$ | $\mathbf{< 10^{-16}}$ |
| GLS | $\mathbf{< 10^{-16}}$ | $\mathbf{1.2 \times 10^{-14}}$ | $\mathbf{< 10^{-16}}$ | $\mathbf{< 10^{-16}}$ |

**Supplementary Table 7: Disease group pairwise comparisons of *visionECG*-derived phenotypes**. Benjamini-Hochberg corrected Mann-Whitney $P$-values are reported for each comparison. $P$-values are bold when $P < 0.05$. DCM, dilated cardiomyopathy; HCM, hypertrophic cardiomyopathy; HF, heart failure; LVM, left ventricular mass; LVEDV, left ventricular end-diastolic volume; LVESV, left ventricular end-systolic volume; LVEDVi, left ventricular end-diastolic volume index; LVESVi, left ventricular end-systolic volume index; LVEF, left ventricular ejection fraction; WT, wall thickness; ES, end systole; ED, end diastole; MAPSE, mitral annular plane systolic excursion; S/L, septal-to-lateral ratio; GCS, global circumferential strain; GLS, global longitudinal strain.

| Feature | DCM & Hypertension | HCM & DCM | HCM & Hypertension | DCM & HF | HCM & HF | HF & Hypertension |
|---|---|---|---|---|---|---|
| LVM | $\mathbf{3.2 \times 10^{-5}}$ | 0.6408 | **0.0014** | 0.8176 | 0.8791 | $\mathbf{3.6 \times 10^{-16}}$ |
| LVEDV | $\mathbf{1.9 \times 10^{-5}}$ | 0.3547 | **0.0024** | 0.6233 | 0.9517 | $\mathbf{< 10^{-16}}$ |
| LVESV | $\mathbf{4.2 \times 10^{-10}}$ | **0.0142** | $\mathbf{4.5 \times 10^{-4}}$ | 0.2786 | 0.5744 | $\mathbf{< 10^{-16}}$ |
| LVEDVi | $\mathbf{2.3 \times 10^{-5}}$ | 0.9530 | $\mathbf{1.3 \times 10^{-5}}$ | 0.6408 | 0.4248 | $\mathbf{< 10^{-16}}$ |
| LVESVi | $\mathbf{1.8 \times 10^{-12}}$ | **0.0204** | $\mathbf{1.6 \times 10^{-6}}$ | 0.1068 | 0.8238 | $\mathbf{< 10^{-16}}$ |
| LVEF | $\mathbf{2.8 \times 10^{-12}}$ | $\mathbf{7.0 \times 10^{-4}}$ | **0.0024** | 0.2786 | 0.1839 | $\mathbf{< 10^{-16}}$ |
| WT Max ES | **0.0248** | 0.7445 | **0.0019** | 0.9594 | 0.5942 | $\mathbf{1.4 \times 10^{-9}}$ |
| WT Max ED | **0.0038** | 0.7536 | **0.0119** | 0.9652 | 0.9594 | $\mathbf{1.1 \times 10^{-9}}$ |
| WT Med ED | **0.0021** | 0.9594 | **0.0074** | 0.9594 | 0.6408 | $\mathbf{1.0 \times 10^{-8}}$ |
| MAPSE | $\mathbf{2.6 \times 10^{-8}}$ | **0.0035** | **0.0065** | 0.7986 | 0.3043 | $\mathbf{< 10^{-16}}$ |
| Sphericity | $\mathbf{6.5 \times 10^{-7}}$ | **0.0040** | **0.0060** | 0.6947 | 0.1075 | $\mathbf{< 10^{-16}}$ |
| S/L | $\mathbf{3.9 \times 10^{-8}}$ | **0.0164** | $\mathbf{3.1 \times 10^{-4}}$ | 0.2816 | 0.6408 | $\mathbf{< 10^{-16}}$ |
| GCS | $\mathbf{3.4 \times 10^{-10}}$ | $\mathbf{6.6 \times 10^{-4}}$ | **0.0143** | 0.2728 | 0.2736 | $\mathbf{< 10^{-16}}$ |
| GLS | $\mathbf{4.3 \times 10^{-11}}$ | **0.0019** | **0.0025** | 0.2728 | 0.2816 | $\mathbf{< 10^{-16}}$ |

**Supplementary Table 8: Abnormal phenotype discrimination across four models**. Area under receiver operating characteristic curve (AUC), sensitivity and specificity at the Youden-optimal threshold with 95% confidence intervals for per task are reported. The bottom row of each non-reference model reports the two-sided DeLong $P$-value for the AUC difference against *visionECG*. $P$-values are bold when $P < 0.05$. LVEF, left ventricular ejection fraction; LVEDVi, left ventricular end-diastolic volume index; WT, wall thickness.

| Model | | LVEF≤45 (%) | LVEF≤50 (%) | LVEDVi F≥62 (ml/m$^2$) | LVEDVi M≥75 (ml/m$^2$) | Max WT≥13 (mm) | Max WT≥15 (mm) |
|---|---|---|---|---|---|---|---|
| ECG Parameters | AUC | 0.76 [0.74, 0.79] | 0.73 [0.71, 0.75] | 0.69 [0.68, 0.70] | 0.72 [0.70, 0.73] | 0.64 [0.63, 0.65] | 0.75 [0.73, 0.77] |
| | Sensitivity | 0.61 [0.56, 0.67] | 0.48 [0.45, 0.51] | 0.66 [0.64, 0.67] | 0.61 [0.60, 0.63] | 0.55 [0.53, 0.56] | 0.64 [0.59, 0.67] |
| | Specificity | 0.79 [0.79, 0.80] | 0.83 [0.82, 0.83] | 0.63 [0.60, 0.65] | 0.71 [0.69, 0.72] | 0.69 [0.68, 0.69] | 0.73 [0.73, 0.74] |
| | DeLong | **0.014** | **$< 10^{-16}$** | **$< 10^{-16}$** | **$< 10^{-16}$** | **$< 10^{-16}$** | **$< 10^{-16}$** |
| EchoNext-Mini | AUC | 0.73 [0.70, 0.76] | 0.71 [0.69, 0.73] | – | – | 0.63 [0.62, 0.64] | 0.73 [0.71, 0.75] |
| | Sensitivity | 0.60 [0.55, 0.65] | 0.61 [0.57, 0.64] | – | – | 0.58 [0.56, 0.59] | 0.74 [0.69, 0.78] |
| | Specificity | 0.77 [0.76, 0.77] | 0.68 [0.68, 0.69] | – | – | 0.61 [0.60, 0.62] | 0.58 [0.57, 0.58] |
| | DeLong | **$1.0 \times 10^{-8}$** | **$< 10^{-16}$** | – | – | **$< 10^{-16}$** | **$< 10^{-16}$** |
| EchoNext-Mini Re-trained Classifier | AUC | 0.73 [0.70, 0.76] | 0.72 [0.70, 0.74] | 0.70 [0.69, 0.72] | 0.73 [0.71, 0.74] | 0.72 [0.71, 0.73] | 0.76 [0.74, 0.78] |
| | Sensitivity | 0.50 [0.45, 0.55] | 0.47 [0.44, 0.50] | 0.65 [0.64, 0.66] | 0.66 [0.64, 0.67] | 0.70 [0.68, 0.71] | 0.62 [0.57, 0.66] |
| | Specificity | 0.88 [0.87, 0.88] | 0.86 [0.85, 0.86] | 0.63 [0.61, 0.66] | 0.66 [0.64, 0.67] | 0.67 [0.66, 0.67] | 0.76 [0.75, 0.76] |
| | DeLong | **$7.1 \times 10^{-7}$** | **$< 10^{-16}$** | **$< 10^{-16}$** | **$< 10^{-16}$** | **$< 10^{-16}$** | **$< 10^{-16}$** |
| *visionECG* Phenotypes | AUC | 0.80 [0.77, 0.83] | 0.82 [0.80, 0.83] | 0.83 [0.82, 0.84] | 0.82 [0.81, 0.83] | 0.80 [0.79, 0.80] | 0.90 [0.89, 0.92] |
| | Sensitivity | 0.58 [0.53, 0.63] | 0.65 [0.62, 0.68] | 0.76 [0.74, 0.77] | 0.74 [0.73, 0.75] | 0.70 [0.68, 0.71] | 0.79 [0.75, 0.82] |
| | Specificity | 0.90 [0.89, 0.90] | 0.83 [0.83, 0.84] | 0.76 [0.74, 0.78] | 0.72 [0.70, 0.73] | 0.74 [0.73, 0.75] | 0.86 [0.86, 0.87] |

**Supplementary Table 9: Abnormal phenotype discrimination on the external test cohort.** Area under the receiver operating characteristic curve (AUC), sensitivity and specificity are reported with 95% bootstrap confidence intervals for the external test set (n=5000, 2,500 per validation and test splits of the dataset). Sensitivity and specificity were calculated at the Youden-optimal threshold. LVEF, left ventricular ejection fraction; WT, wall thickness.

| Abnormality | LVEF≤35 (%) | LVEF≤45 (%) | LVEF≤50 (%) | Max WT≥13 (mm) | Max WT≥15 (mm) | Max WT≥16 (mm) |
|---|---|---|---|---|---|---|
| AUC | 0.81 [0.79, 0.84] | 0.80 [0.78, 0.82] | 0.79 [0.77, 0.81] | 0.68 [0.65, 0.70] | 0.67 [0.63, 0.71] | 0.70 [0.65, 0.75] |
| Sensitivity | 0.79 [0.75, 0.83] | 0.83 [0.80, 0.86] | 0.80 [0.77, 0.83] | 0.71 [0.67, 0.75] | 0.70 [0.63, 0.77] | 0.76 [0.67, 0.83] |
| Specificity | 0.71 [0.69, 0.73] | 0.61 [0.59, 0.63] | 0.63 [0.61, 0.65] | 0.53 [0.51, 0.55] | 0.49 [0.47, 0.51] | 0.48 [0.46, 0.50] |

**Supplementary Table 10: Disease prediction across four models**. Area under receiver operating characteristic curve (AUC), sensitivity and specificity at the Youden-optimal threshold with 95% bootstrap confidence intervals for per task are reported. The bottom row of each non-reference model reports the two-sided DeLong $P$-value for the AUC difference against *visionECG*. $P$-values are bold when $P < 0.05$. CM, cardiomyopathy; HCM, hypertrophic cardiomyopathy; DCM, dilated cardiomyopathy; HF, heart failure; MI, myocardial infarction.

| Model | | Cardiac Disease | HF | MI | CM | HCM | DCM |
|---|---|---|---|---|---|---|---|
| *visionECG* LVEF & Max WT | AUC | 0.72 [0.69, 0.74] | 0.78 [0.73, 0.82] | 0.69 [0.66, 0.71] | 0.75 [0.62, 0.84] | 0.61 [0.42, 0.75] | 0.89 [0.77, 0.95] |
| | Sensitivity | 0.70 [0.68, 0.71] | 0.69 [0.60, 0.77] | 0.63 [0.58, 0.68] | 0.52 [0.32, 0.70] | 0.23 [0.00, 0.50] | 0.79 [0.50, 0.94] |
| | Specificity | 0.63 [0.59, 0.67] | 0.70 [0.69, 0.72] | 0.67 [0.65, 0.69] | 0.82 [0.81, 0.83] | 0.93 [0.92, 0.93] | 0.85 [0.84, 0.86] |
| | DeLong | **$< 10^{-16}$** | **0.035** | **$< 10^{-16}$** | **0.036** | **$3.3 \times 10^{-5}$** | 0.41 |
| ECG Parameters | AUC | 0.75 [0.73, 0.78] | 0.77 [0.72, 0.81] | 0.73 [0.70, 0.75] | 0.84 [0.73, 0.91] | 0.71 [0.48, 0.83] | 0.89 [0.75, 0.95] |
| | Sensitivity | 0.84 [0.82, 0.85] | 0.68 [0.60, 0.76] | 0.52 [0.47, 0.57] | 0.70 [0.50, 0.84] | 0.31 [0.06, 0.62] | 0.64 [0.32, 0.87] |
| | Specificity | 0.50 [0.46, 0.55] | 0.76 [0.74, 0.77] | 0.79 [0.78, 0.81] | 0.80 [0.78, 0.81] | 0.92 [0.91, 0.93] | 0.93 [0.92, 0.94] |
| | DeLong | **$2.1 \times 10^{-11}$** | **0.0082** | **$1.6 \times 10^{-9}$** | 0.95 | **0.0023** | 0.70 |
| EchoNext-Mini Re-trained Classifier | AUC | 0.75 [0.73, 0.77] | 0.77 [0.73, 0.82] | 0.74 [0.71, 0.76] | 0.81 [0.68, 0.88] | 0.67 [0.47, 0.82] | 0.88 [0.81, 0.92] |
| | Sensitivity | 0.74 [0.72, 0.76] | 0.51 [0.43, 0.59] | 0.62 [0.57, 0.67] | 0.59 [0.40, 0.76] | 0.69 [0.33, 0.91] | 0.79 [0.49, 0.94] |
| | Specificity | 0.64 [0.60, 0.68] | 0.86 [0.85, 0.87] | 0.74 [0.73, 0.76] | 0.87 [0.86, 0.88] | 0.58 [0.56, 0.59] | 0.86 [0.85, 0.87] |
| | DeLong | **$7.4 \times 10^{-10}$** | **0.047** | **$1.8 \times 10^{-6}$** | 0.48 | **$5.8 \times 10^{-4}$** | 0.57 |
| *visionECG* Phenotypes | AUC | 0.83 [0.81, 0.85] | 0.82 [0.78, 0.86] | 0.81 [0.79, 0.83] | 0.85 [0.75, 0.90] | 0.92 [0.86, 0.95] | 0.91 [0.81, 0.95] |
| | Sensitivity | 0.77 [0.76, 0.78] | 0.71 [0.63, 0.78] | 0.78 [0.73, 0.82] | 0.56 [0.33, 0.73] | 0.92 [0.57, 1.00] | 0.36 [0.14, 0.64] |
| | Specificity | 0.72 [0.68, 0.77] | 0.77 [0.76, 0.78] | 0.71 [0.69, 0.72] | 0.89 [0.88, 0.90] | 0.75 [0.74, 0.77] | 0.97 [0.96, 0.98] |

**Supplementary Table 11: Event summary for the longitudinal cardiovascular risk stratification**. The Event column reports participants whose event was observed during follow-up. Time to Event column is the interval from ECG acquisition to the first recorded event, summarised as the median [Q1, Q3] among participants with an observed event. HF, heart failure; MI, myocardial infarction; MACE, major adverse cardiovascular events; mo, months.

| Disease | Event n (%) | Time to Event (mo) |
|---|---|---|
| HF | 258 (1.47%) | 25.68 [13.79, 44.46] |
| MI | 530 (3.02%) | 23.52 [13.83, 39.06] |
| MACE | 803 (4.57%) | 23.79 [13.29, 40.76] |

**Supplementary Table 12: Cardiovascular risk stratification with Cox proportional-hazard models.** Harrell's C-index and hazard ratios per one standard deviation (SD) of two feature groups, *visionECG* phenotypes and ECG parameters, are reported with 95% confidence intervals. $P$ values are calculated using the Wald test. $P$-values are bold when $P < 0.05$. HF, heart failure; MI, myocardial infarction; MACE, major adverse cardiovascular events.

| Disease | Composite Risk Probability | C-index (95% CI) | Hazard Ratio per SD (95% CI) | Hazard Ratio $P$ |
|---|---|---|---|---|
| HF | ECG parameters | 0.628 [0.543, 0.713] | 1.480 [1.330, 1.647] | $\mathbf{6.7 \times 10^{-13}}$ |
| | *visionECG* phenotypes | 0.762 [0.700, 0.824] | 1.790 [1.600, 2.003] | $\mathbf{< 10^{-16}}$ |
| MI | ECG parameters | 0.508 [0.447, 0.568] | 1.045 [0.862, 1.266] | 0.6538 |
| | *visionECG* phenotypes | 0.563 [0.509, 0.618] | 1.254 [1.028, 1.530] | **0.0259** |
| MACE | ECG parameters | 0.565 [0.516, 0.614] | 1.181 [1.088, 1.281] | $\mathbf{1.0 \times 10^{-4}}$ |
| | *visionECG* phenotypes | 0.615 [0.569, 0.660] | 1.290 [1.179, 1.412] | $\mathbf{2.8 \times 10^{-8}}$ |

**Supplementary Table 13: Likelihood-ratio tests of nested Cox proportional-hazards models.** The full model included both feature groups, *visionECG* phenotypes and ECG parameters. Each reduced model included only one feature group. Likelihood-ratio tests compared each reduced model with the full model. HF, heart failure; MI, myocardial infarction; MACE, major adverse cardiovascular events.

| Disease | Feature Group | C-index (95% CI) | Feature Group Removal Tests $P$ |
|---|---|---|---|
| HF | ECG & *visionECG* | 0.752 [0.685, 0.818] | — |
| | ECG only | 0.628 [0.543, 0.713] | $\mathbf{7.2 \times 10^{-10}}$ |
| | *visionECG* only | 0.762 [0.700, 0.824] | 0.3708 |
| MI | ECG & *visionECG* | 0.566 [0.513, 0.620] | — |
| | ECG only | 0.508 [0.447, 0.568] | **0.0272** |
| | *visionECG* only | 0.563 [0.509, 0.618] | 0.8657 |
| MACE | ECG & *visionECG* | 0.606 [0.559, 0.652] | — |
| | ECG only | 0.565 [0.516, 0.614] | $\mathbf{2.0 \times 10^{-4}}$ |
| | *visionECG* only | 0.615 [0.569, 0.660] | 0.0891 |

**Supplementary Table 14: UKB data-field IDs and ICD-10 codes used to define the disease and reference groups.** ICD-10, the tenth revision of the International Classification of Diseases; UKB, UK Biobank; HF, heart failure; MI, myocardial infarction; HCM, hypertrophic cardiomyopathy; DCM, dilated cardiomyopathy; MACE, major adverse cardiovascular events; BMI, body mass index.

| Group | Definition | UKB data-field ID(s) | ICD-10 code(s) |
|---|---|---|---|
| HF | Diagnosis of heart failure | 40001, 40002, 41270 | I11.0, I13.0, I13.2, I50 |
| MI | Diagnosis of myocardial infarction | 40001, 40002, 41270 | I21–I23 |
| HCM | Diagnosis of hypertrophic cardiomyopathy | 40001, 40002, 41270 | I42.1, I42.2 |
| DCM | Diagnosis of dilated cardiomyopathy | 40001, 40002, 41270 | I42.0, I42.6, I42.7 |
| Hypertension | Diagnosed hypertension | 40001, 40002, 41270, 1065, 1072, 1073 | I10, I11.0, I11.9, I12.0, I12.9, I13.0, I13.1, I13.2, I13.9, I15.0, I15.1, I15.2, I15.9 |
| MACE | Composite of MI, HF, cardiac arrest, stroke, or cardiovascular death | 40001, 40002, 41270 | I11.0, I13.0, I13.2, I21–I23, I46, I50, I60–I64 |
| Control | Participants without recorded cardiac disease | Refer to Supplementary Table 15 | Refer to Supplementary Table 15 |
| Healthy | Participants without recorded cardiac disease, chronic respiratory disease, hypercholesterolaemia, or diabetes mellitus, and with BMI $< 30\,\mathrm{kg/m^2}$ | Refer to Supplementary Table 15 | Refer to Supplementary Table 15 |

**Supplementary Table 15: Diagnostic codes used to define study groups in UKB**. ICD-10, the tenth revision of the International Classification of Diseases, a system used by healthcare providers to code and classify diseases and health conditions. ICD-9, the ninth revision of the International Classification of Diseases.

| Disease status | ICD-10 Code(s) | ICD-9 Code(s) | UKB data-field ID(s) |
|---|---|---|---|
| Hypercholesterolaemia | E78 | 27202, 27209, 27200, 2720 | 1473 |
| Diabetes mellitus | E10-E14 | | 2443, 1220, 1221, 1222, 1223, 1276, 1468, 1607 |
| General presence of cardiac disease | I00-I09, I20-I25, I30-I52 | | |
| Chronic respiratory disease | J40, J410, J411, J418, J42, J431, J432, J438, J439, J440, J441, J448, J449, J450, J451, J458, J459, J46, J47 | 4241 | 1490, 1586 |

### Supplementary movies

**Supplementary Movie 1. Dynamic visualisation of cardiac phenotype measurements**. Measurements of cardiac volumetric, geometric and motion-derived phenotypes are dynamically visualised on *visionECG*-generated mesh sequences throughout the cardiac cycle. a, radial measurements and wall thickness; b, circumferential measurements and circumferential strain; c, longitudinal measurements and longitudinal strain; d, MAPSE, sphericity and volume measurements. Animated version of Figure 3.

**Supplementary Movie 2. Dynamic visualisation of case-specific *visionECG*-generated meshes**. ECG waveforms are shown with their corresponding *visionECG*-generated left ventricular mesh sequences, dynamically visualised throughout the cardiac cycle. a is the healthy group average; b, a case of hypertrophic cardiomyopathy (HCM); c, a case of dilated cardiomyopathy (DCM); d, a case of hypertension.
